# Human influenza virus infection elicits distinct patterns of monocyte and dendritic cell mobilization in blood and the nasopharynx

**DOI:** 10.1101/2022.01.18.22269508

**Authors:** S. Vangeti, S. Falck-Jones, M. Yu, B. Österberg, S. Liu, M. Asghar, K. Sondén, J. Albert, N. Johansson, A. Färnert, A. Smed-Sörensen

## Abstract

During respiratory viral infections, the precise roles of monocytes and dendritic cells (DCs) in the nasopharynx in limiting infection and influencing disease severity are incompletely described. We studied circulating and nasopharyngeal monocytes and DCs in healthy individuals and in patients with mild respiratory infections (primarily influenza A virus, IAV). As compared to healthy controls (HCs), patients with acute IAV infection displayed reduced DC but increased intermediate monocytes frequencies in blood, and an accumulation of most monocyte and DC subsets in the nasopharynx. IAV patients had more mature monocytes and DCs in the nasopharynx, and higher levels of TNFα, IL-6 and IFNα in plasma and the nasopharynx. In blood, monocytes, the most frequent cellular source of TNFα during IAV infection, remained responsive to additional stimulation with TLR7/8L. Immune responses in older patients skewed towards increased monocytes rather than DCs suggesting a contributory role for monocytes in disease severity. In patients with other respiratory virus infections, we observed changes in monocyte and DC frequencies in the nasopharynx distinct from IAV patients, while differences in blood were more similar across patient groups. Together, our findings demonstrate tissue-specific and pathogen-specific patterns of monocyte and DC function during human respiratory viral infections and highlight the importance of comparative investigations in blood and the nasopharynx.

## INTRODUCTION

Respiratory viral infections cause significant global disease burden with influenza, or flu, being responsible for a significant portion. An estimated 1 billion cases of influenza occur annually resulting in approximately 3-5 million severe cases and 290,000-650,000 deaths (1). In addition to seasonal epidemics caused by both influenza A or B virus (IAV and IBV, respectively), IAV can also cause pandemics. The majority of infections remain asymptomatic or develop mild to moderate respiratory disease, characterized by fever, nasal congestion, cough, and muscle aches. Severe disease mainly affects infants, pregnant women, and the elderly or immunocompromised, but can also occur in otherwise healthy individuals (2, 3). The determinants of disease severity are still incompletely understood but may include properties of the virus, environmental factors, genetics, and immune responses of the patient (4, 5). Other agents causing an influenza-like illness during influenza season include respiratory syncytial virus (RSV) and seasonal coronaviruses (OC43, HKU1, 229E, and NL63). Similar to IAV, coronaviruses are capable of causing pandemics, most notably the ongoing coronavirus disease 2019 (COVID-19) caused by the severe acute respiratory syndrome coronavirus 2 (SARS-CoV- 2) (6).

IAV is primarily transmitted via inhalation of virus-containing aerosols or droplets, and mainly targets respiratory epithelial cells (7, 8), with the nasopharynx being the initial site of virus replication. IAV generally remains localized to the airways, despite signs of systemic inflammation (9). At the site of infection, resident innate immune cells including monocytes and dendritic cells (DCs) rapidly respond to the presence of virus by secreting cytokines, interferons, and chemokines to limit viral spread and recruit immune cells (10, 11). Monocytes and DCs shape the specificity and strength of the subsequent adaptive responses (12, 13). In blood, three subsets of monocytes are found: the CD14+CD16– classical monocytes (CMs), the most frequent subset at steady state, and the further differentiated CD14+CD16+ intermediate monocytes (IMs) and CD14–CD16+ nonclassical monocytes (NCMs) (14–16). Blood IMs expand rapidly in response to inflammation, infection, or vaccination (3, 12, 17). In addition, monocytes extravasate to tissue where they play an important role in innate immune protection. Monocytes also secrete TNFα, a major regulator of innate immune function that is central to the cytokine storm associated with IAV infection (18). Of the DCs, the CD1c+ myeloid DCs (MDCs) excel at activating naïve T cells (19); the CD141+ MDCs can cross-present antigens via MHC-I (20); and the CD123+ plasmacytoid DCs (PDCs) mediate type I IFN responses (21). Monocytes and DCs vary in distribution and function, depending on the anatomical compartment (22, 23). Moreover, monocytes and DCs are susceptible to IAV infection in vitro, and the cytopathic nature of the virus may impair their antigen processing and presenting functions (24–26), delaying recovery and normalization of immune cell distribution and function (27, 28).

Studies have shown that monocytes and DCs are recruited to the nasopharynx following infection with 2009 H1N1pdm IAV strains (3), and in individuals hospitalized with severe influenza infections (29, 30). Disease severity in hospitalized patients has been shown to correlate with (i) monocyte recruitment and increased levels of MCP3, IFNα-2 and IL-10 in the nasal compartment (3) and (ii) strong TNF-producing monocytic responses in blood (2) and inflammatory, neutrophil-dominant patterns (31). Despite accounting for a comparatively greater burden of disease, immune responses during mild seasonal influenza infections remain less studied. Therefore, the roles played by monocytes and DCs in contributing to or mitigating mild influenza disease are largely unknown. Additionally, while the response of blood monocytes and DCs to IAV has been studied well in vitro and in animal models (24, 25, 27, 32- 35), few studies compare responses between blood and the nasopharynx in human infections (31, 32, 36). Immune cell behaviour in the nasopharynx during IBV and RSV infections has not been studied in great detail but evidence of DC mobilization to the nasal cavity has been reported (29). Studies on immune responses to mild SARS-CoV-2 infection have also primarily focused on blood and rarely the upper airways (37).

Here, we determined monocyte and DC subset distribution, maturation and function in both blood and, for the first time, the nasopharynx, in patients with mild to moderate seasonal influenza and influenza-like infections. The methods described in this study allowed us to investigate airway immunity in a larger cohort of patients with SARS-CoV-2 infection (38, 39), showing that methods to study the immune responses in the nasopharynx during acute disease are essential tools as we face the possibility of future pandemics. Comparing the dynamics of systemic and nasopharyngeal immune function will add to our understanding of the roles of monocytes and DCs in shaping the nature and magnitude of inflammation and subsequently disease severity during respiratory viral infections.

## RESULTS

### Study subject characteristics

During three consecutive influenza seasons (2016–2018), 84 adults with symptoms of influenza-like illness (ILI) were included in the study. Blood, nasal swabs and nasopharyngeal aspirates were collected (Figure 1A). IAV infection was confirmed by PCR in 40 patients while 44 patients had other infections (IBV: 10, RSV: 6, other viruses: 2, bacteria: 3 and unknown aetiology: 23) despite presenting with similar symptoms (Figure 1B), consistent with inclusion based on ILI. Patients with bacterial infection, other viral infection or infections of unknown etiology were excluded from further analysis leaving 56 patients with confirmed IAV, IBV or RSV infection. The severity of disease in patients was categorized as “mild” or “moderate” (detailed description in Methods). Of the 56 IAV/IBV/RSV patients, 8 had moderately severe disease and 21 were hospitalized (Table 1). During the COVID-19 pandemic in 2020, we included 8 adults with PCR+ SARS-CoV-2 infection (7 with mild and 1 with moderate disease) for this study based on the availability of paired blood and NPA samples from the time of inclusion. Sixteen healthy controls (HCs) were included and sampled identically as patients.

**Figure 1.**
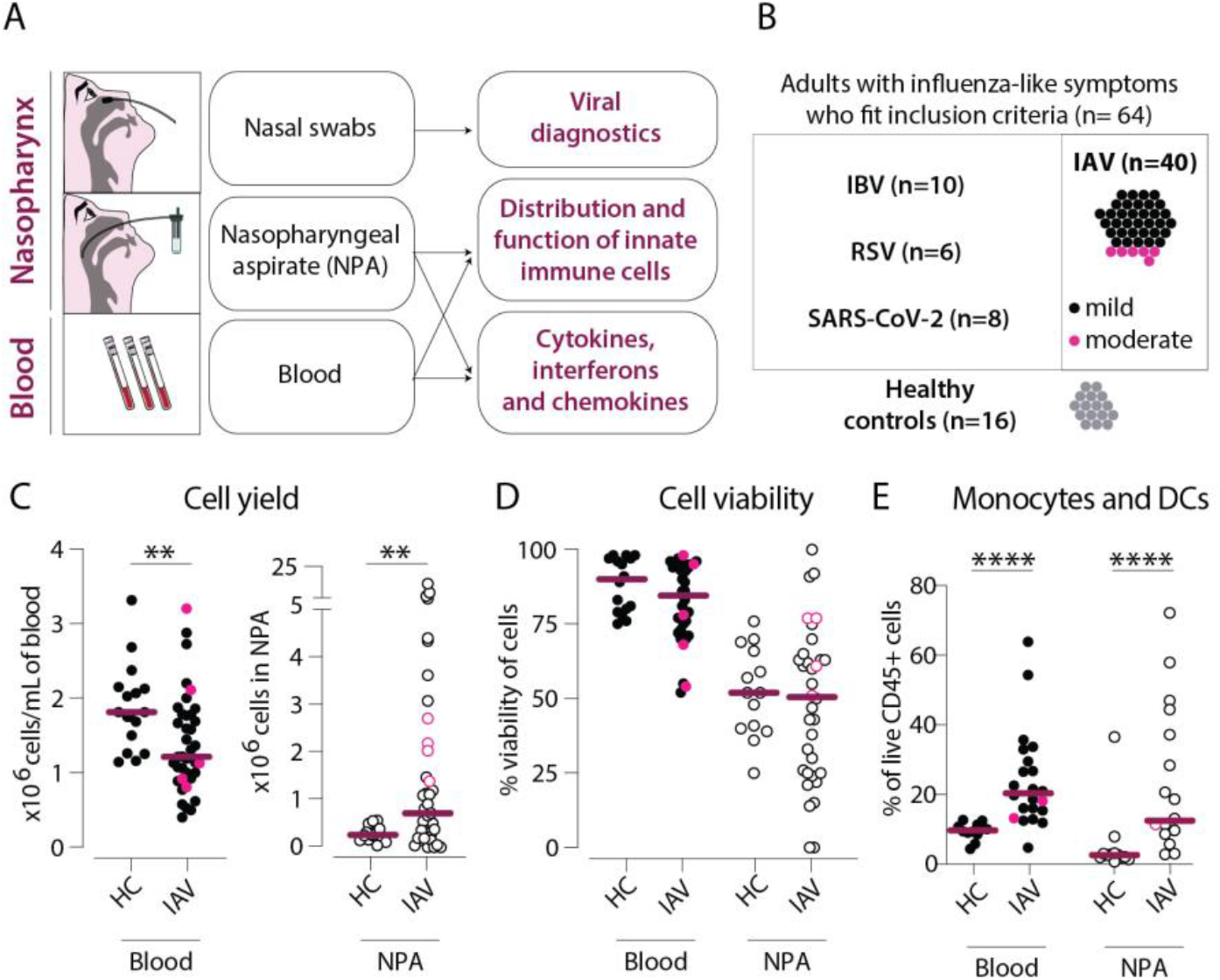
Cellular infiltration into the nasopharynx during IAV infection is largely due to accumulation of lin–HLA-DR+ cells. (**A**) Nasal swabs, nasopharyngeal aspirates (NPA), and peripheral blood samples were collected from patients with acute symptoms of influenza-like symptoms and during their convalescence as well as from healthy controls (HCs). (**B**) 40 of 64 patients with influenza-like symptoms were confirmed to be infected with influenza A virus (IAV) by PCR and were included in the study. Immunological analyses were performed on acute (n=40) and convalescent (n=11) samples from the IAV patients and HC (n=16). IAV patients with mild disease, as defined by peak respiratory SOFA or mSOFA score of 1 or 2 are indicated in black and those with moderate disease (mSOFA score of 3 or 4) are indicated in pink. (**C-E**) Scatter plots show data from individual subjects and lines indicate median values. Patients with mild disease are indicated in black and those with moderate disease are indicated in pink. (**C**) x10^6^ PBMCs (per mL blood, filled circles) and x10^6^ total NPA cells (open circles) obtained from IAV patients and HCs. (**D**) Cell viability of PBMCs and NPA from patients and HCs was assessed using trypan blue exclusion staining and manual counting. (**E**) Frequency of lin–HLA-DR+ cells (monocytes and myeloid dendritic cells) of live CD45+ cells in blood and NPA from HCs (n=16) and IAV patients (n=22). Differences between IAV patients and HCs were assessed using Mann-Whitney test and considered significant at p<0.05 (**p<0.01, ****p<0.0001).

**Table 1.**
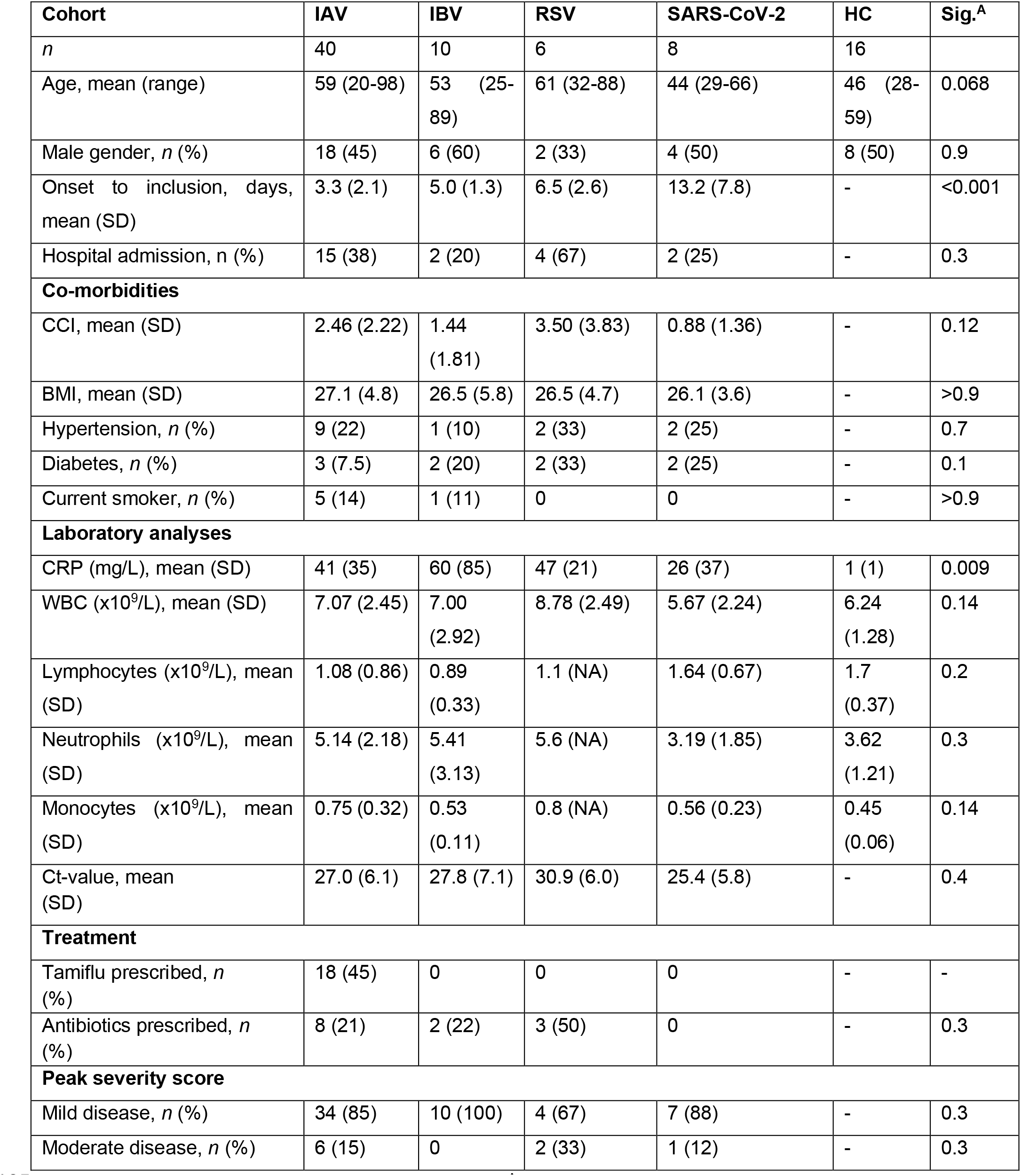
Patient and control characteristics. ^A^Statistical significance was determined by 1-way ANOVA, Fisher’s exact test, or Pearson’s χ2 test. CCI: Charlson Comorbidity Index. BMI: Body Mass Index. CRP: C-reactive protein. WBC: White Blood Cells. Ct: Cycle threshold. Normal range: BMI: 18.5 to 24.9, CRP <3 mg/L, WBC 3.5 × 10^9^/L to 8.8 × 10^9^/L, lymphocytes 1.1 × 10^9^/L to 3.5 × 10^9^/L, neutrophils 1.6 × 10^9^/L to 5.9 × 10^9^/L, monocytes 0.2 × 10^9^/L to 0.8 × 10^9^/L.

| Cohort | IAV | IBV | RSV | SARS-CoV-2 | HC | Sig.<sup>A</sup> |
| --- | --- | --- | --- | --- | --- | --- |
| *n* | 40 | 10 | 6 | 8 | 16 |  |
| Age, mean (range) | 59 (20-98) | 53 (25-89) | 61 (32-88) | 44 (29-66) | 46 (28-59) | 0.068 |
| Male gender, *n* (%) | 18 (45) | 6 (60) | 2 (33) | 4 (50) | 8 (50) | 0.9 |
| Onset to inclusion, days, mean (SD) | 3.3 (2.1) | 5.0 (1.3) | 6.5 (2.6) | 13.2 (7.8) | - | <0.001 |
| Hospital admission, *n* (%) | 15 (38) | 2 (20) | 4 (67) | 2 (25) | - | 0.3 |
| **Co-morbidities** |  |  |  |  |  |  |
| CCI, mean (SD) | 2.46 (2.22) | 1.44 (1.81) | 3.50 (3.83) | 0.88 (1.36) | - | 0.12 |
| BMI, mean (SD) | 27.1 (4.8) | 26.5 (5.8) | 26.5 (4.7) | 26.1 (3.6) | - | >0.9 |
| Hypertension, *n* (%) | 9 (22) | 1 (10) | 2 (33) | 2 (25) | - | 0.7 |
| Diabetes, *n* (%) | 3 (7.5) | 2 (20) | 2 (33) | 2 (25) | - | 0.1 |
| Current smoker, *n* (%) | 5 (14) | 1 (11) | 0 | 0 | - | >0.9 |
| **Laboratory analyses** |  |  |  |  |  |  |
| CRP (mg/L), mean (SD) | 41 (35) | 60 (85) | 47 (21) | 26 (37) | 1 (1) | 0.009 |
| WBC (x10<sup>9</sup>/L), mean (SD) | 7.07 (2.45) | 7.00 (2.92) | 8.78 (2.49) | 5.67 (2.24) | 6.24 (1.28) | 0.14 |
| Lymphocytes (x10<sup>9</sup>/L), mean (SD) | 1.08 (0.86) | 0.89 (0.33) | 1.1 (NA) | 1.64 (0.67) | 1.7 (0.37) | 0.2 |
| Neutrophils (x10<sup>9</sup>/L), mean (SD) | 5.14 (2.18) | 5.41 (3.13) | 5.6 (NA) | 3.19 (1.85) | 3.62 (1.21) | 0.3 |
| Monocytes (x10<sup>9</sup>/L), mean (SD) | 0.75 (0.32) | 0.53 (0.11) | 0.8 (NA) | 0.56 (0.23) | 0.45 (0.06) | 0.14 |
| Ct-value, mean (SD) | 27.0 (6.1) | 27.8 (7.1) | 30.9 (6.0) | 25.4 (5.8) | - | 0.4 |
| **Treatment** |  |  |  |  |  |  |
| Tamiflu prescribed, *n* (%) | 18 (45) | 0 | 0 | 0 | - | - |
| Antibiotics prescribed, *n* (%) | 8 (21) | 2 (22) | 3 (50) | 0 | - | 0.3 |
| **Peak severity score** |  |  |  |  |  |  |
| Mild disease, *n* (%) | 34 (85) | 10 (100) | 4 (67) | 7 (88) | - | 0.3 |
| Moderate disease, *n* (%) | 6 (15) | 0 | 2 (33) | 1 (12) | - | 0.3 |

IAV patients had a mean age of 59 (range: 20-98 years) and on average sought medical attention 3.3 days after the onset of symptoms (SD: 2.1 days) (Table 1). HCs had slightly lower mean age of 46 (range: 28-59 years). IAV patients had a mean Charlson co-morbidity index (CCI) of 2.46 (SD: 2.22), and 36 IAV patients had at least one underlying comorbidity (chronic heart/lung diseases, reduced lung function, kidney insufficiency, diabetes mellitus, asplenia/hyposplenia and malignancies).

### Human IAV infection is characterized by an influx of CD11c+ cells into the nasopharynx

Blood from IAV patients yielded significantly fewer PBMCs/mL compared to HCs (Figure 1C). In contrast, 3-fold higher cell numbers were recovered from the nasopharynx of IAV patients compared to HCs (median 0.77 vs. 0.25 x 10^6^ cells). In fact, 43% of IAV patients had more than 1 x 10^6^ cells recovered from their NPA sample. Viability of PBMCs and NPA cells was variable across individuals, with no statistically significant differences between patients or HCs (Figures 1D). We determined the immune cell distribution in blood and the nasopharynx by flow cytometry (Supplementary Figure 1) on matched PBMC and NPA samples with a minimum 10^5^ cells and ≥70% viability (n=22 IAV patients and n=16 HCs) to obtain high quality data (stained with identical panels and clones of antibodies). The frequencies of live CD45+ immune cells were increased in the NPA of patients as compared to HCs but remained similar in blood between the groups (data not shown). Among the immune cells, we found significantly higher frequencies of lineage (CD3, CD19, CD20, CD56, CD66abce) negative, HLA-DR+ cells, the compartment where monocytes and myeloid DCs can be identified, in both blood (p<0.0001), and the NPA (p<0.01) of IAV patients as compared to HCs (Figure 1E). Therefore, our data show that acute IAV infection results in an influx of monocytes and DCs to the nasopharynx.

**Supplementary Figure 1.**
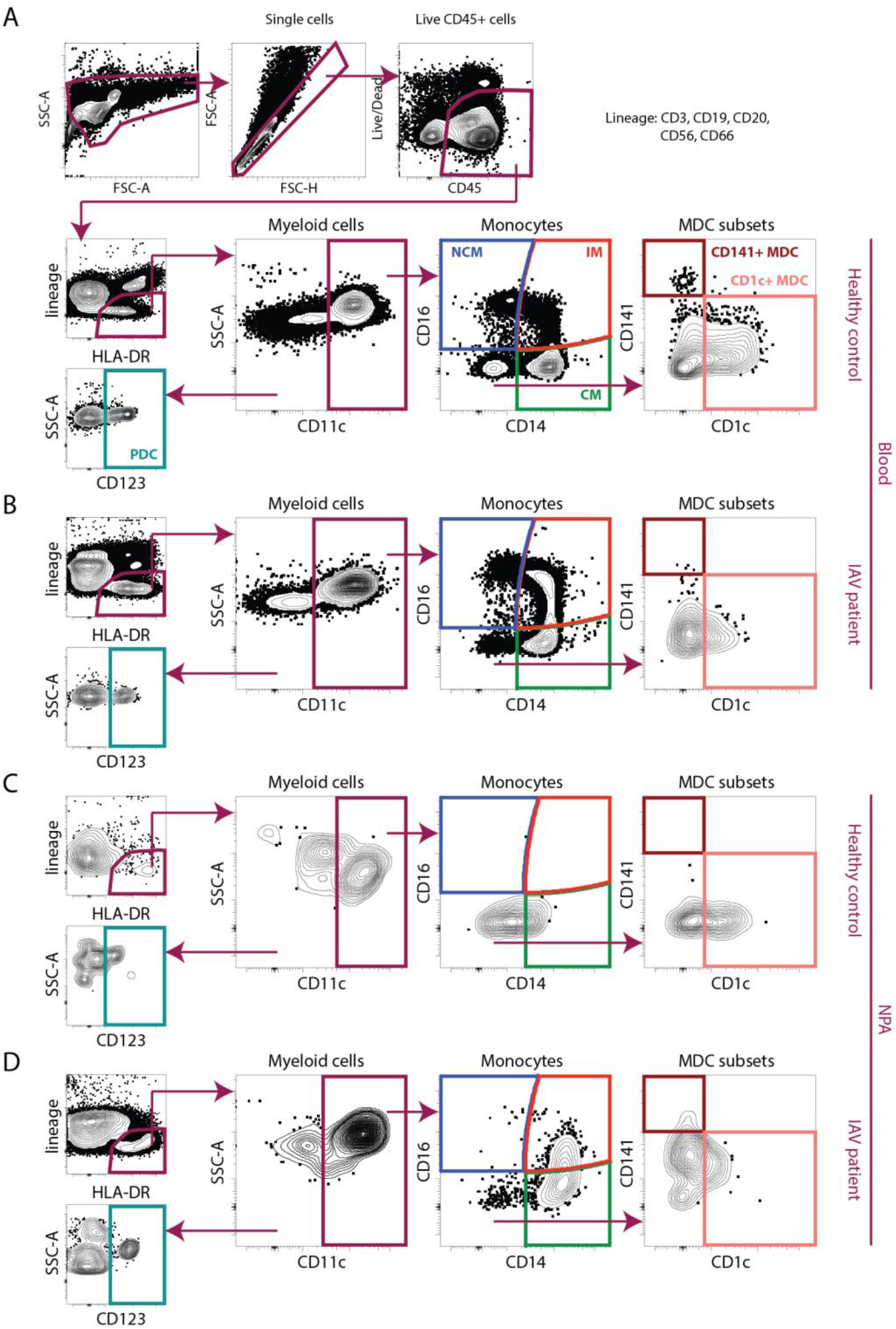
Gating strategy for identification of monocytes and dendritic cells from PBMCs and NPA. Representative sample showing gating on live CD45+ single cells, excluding cells expressing lineage markers (CD3, CD20, CD56 and CD66abce) and identification of HLA-DR expressing cells. From the live CD45+, lineage negative, HLA-DR+ (lin–HLA-DR+) cells, CD11c– cells were gated upon to identify CD123+ plasmacytoid DCs (teal). From the CD11c+ myeloid cells, classical (CD14+CD16−) (green), intermediate (CD14+CD16+) (red) and non-classical (CD14−CD16+) (blue) monocytes were identified. From CD14−CD16− cells, two myeloid DC subsets CD1c+ (coral) and CD141+ (maroon) were identified. Representative samples of using the antibody panel and gating strategy to analyze all 6 subsets (CMs, IMs, NCMs, PDCs, CD1c+ MDCs and CD141+ MDCs) in blood samples from a (**A**) HC and (**B**) IAV patient and NPA samples from a (**C**) HC and (**D**) IAV patient are shown.

### Increased frequencies of classical and intermediate monocytes in the nasopharynx during IAV infection

To identify which monocyte subsets contributed to the changes observed during IAV infection, we analyzed the distribution of the different monocyte subsets (Figure 2A). As expected, CMs were the most frequent monocytes in blood in both patients and HCs, and remained comparable. However, in the nasopharynx of IAV patients as compared to HCs, blood CM frequencies were significantly increased (Figure 2B). Strikingly, IM frequencies were significantly elevated, in both blood and NPA of IAV patients (Figure 2C), while blood NCM appeared to be lower in patients compared to HCs (Figure 2D). Comparing frequencies of monocytes in blood and NPA in the same individual (Figure 2E) illustrates that IAV infection disrupts the pattern of monocyte distribution in the two anatomical compartments seen in healthy individuals- primarily in CMs and IMs. The frequencies of IMs in blood and NPA, and of the CMs in the NPA (solid lines) displayed greatest fluctuation from HCs (dashed line) at fewer days with symptoms (Figures 2F-G). This suggests that changes in the monocyte compartment may occur earlier in the course of infection but these findings should be interpreted with caution since they are based on a single acute phase sample from each patient. In future studies, longitudinal sampling of the same patient would allow for mapping kinetics of monocyte redistribution over the course of acute IAV infection and convalescence. We also compared the frequency of IMs in blood and nasopharynx with the age of IAV patients and found a negative correlation in blood (R=–0.55, p=0.0008) (Figure 2H) but a positive correlation in the nasopharynx (R=0.46, p=0.045) (Figure 2I). In HCs, age and IM frequencies were not significantly correlated in blood or NPA (data not shown). A subset of IAV patients (n=11) returned for sampling during convalescence (≥ 4 weeks after initial sampling). We observed that frequencies of CMs (in blood) and IMs (blood and NPA) in convalescent individuals were reduced and closer to values seen in HCs (Figure 2J). Collectively, we found that the increased immune cell presence in the nasopharynx during acute IAV infection could be, to a large extent, attributed to increased frequencies of IMs as well as CMs which normalized during convalescence. Acute IAV infection resulted in altered monocyte distribution, in particular at the site of infection and was more pronounced in older patients.

**Figure 2.**
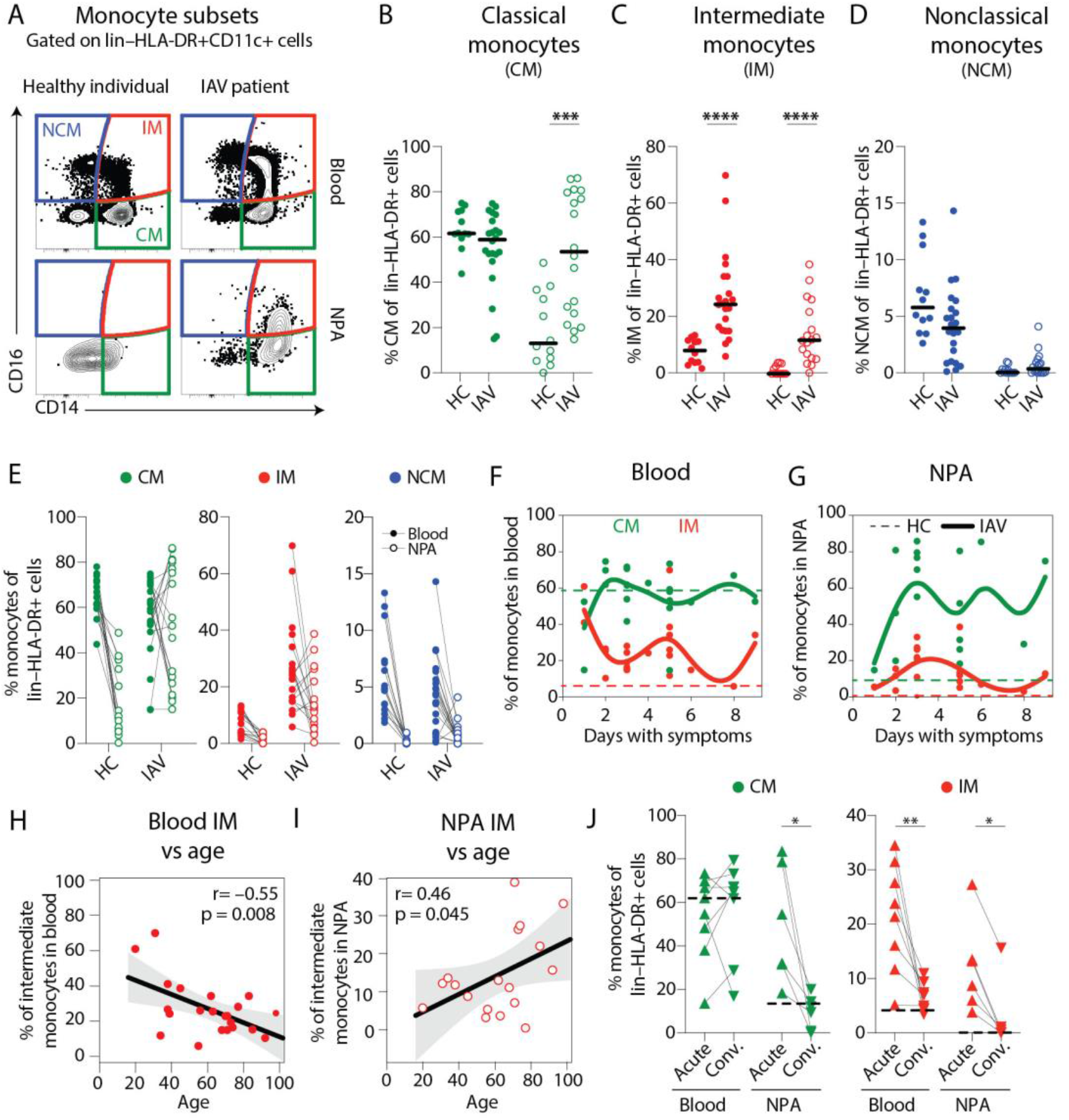
Higher frequencies of intermediate monocytes in blood and the nasopharynx from IAV patients compared to healthy controls. (**A**) Contour plots show CD14+CD16−classical monocytes (CMs, green), CD14+CD16+ intermediate monocytes (IMs, red) and CD14−CD16+ non-classical monocytes (NCMs, blue) in blood (top) and NPA (bottom) from one representative HC and one representative IAV patient determined by flow cytometry. (**B-D**) Scatter plots show frequencies of (**B**) CMs, (**C**) IMs and (**D**) NCMs in PBMCs and NPA from HCs (n=12) and IAV patients (n=22). Graphs show data from individual subjects and lines represent median values. Differences between IAV patients and HCs were assessed using Mann-Whitney test and considered significant at p<0.05 (***p<0.001, ****p<0.0001). (**E**) displays frequencies of monocyte subsets from **B-D** again, with lines connecting values in blood (filled circles) and NPA (open circles) in the same individual. (**F-G**) Temporal changes in CM and IM frequencies as a function of days with symptoms. Solid lines illustrate the trend for CMs (green) and IMs (red) in (**F**) blood and (**G**) NPA of IAV patients as compared to median values observed in HCs (dashed lines). (**H-I**) Scatter plots and line of fit display bivariate linear regression analysis between age and IM frequency in (**H**) blood and (**I**) NPA of IAV patients. The shaded area represents the 95% confidence region for the fitted line. R represents Spearman ρ and differences were considered significant at p<0.05. (**J**) Graphs depict frequencies of CMs (green) and IMs (red) in blood (n=8) and the NPA (n=6) in the acute (upward triangles) and convalescent phase (downward triangles) in IAV patients. Dashed lines depict median frequency values from HCs in blood and NPA. Differences between acute and convalescent phase values were assessed using Wilcoxon matched-pairs signed rank test and considered significant at p<0.05 (*p<0.05 and **p<0.01).

### Dendritic cell subsets infiltrate the nasopharynx during IAV infection

DCs may also account for redistribution in the myeloid cell compartment. In HCs, the nasopharynx was virtually devoid of DCs, and CD1c+ MDCs, CD141+ MDCs and plasmacytoid DCs (PDCs) were only identified in a subset of HCs (Supplementary figure 1 and Figure 3A). In contrast, during acute IAV infection, all DC subsets were significantly increased in the nasopharynx of patients (Figure 3B-D). Meanwhile in blood, DC frequencies decreased significantly in IAV patients compared to HCs (Figures 3B-D). This reciprocal pattern is further illustrated by comparing the DC frequencies in blood and NPA in the same individual (Figure 3E). Looking over days of symptoms, the frequencies of DCs in blood remained lower in IAV patients (solid lines) as compared to HCs (dashed lines) throughout the sampling window (0-9 days with symptoms), with the most striking reduction evident at the earliest times (Figures 3F-G). Conversely in the nasopharynx, DC frequencies in patients fluctuated over time but remained higher than in controls. In contrast to IMs, we found an inverse correlation between age and the frequency of CD1c+ MDCs in NPA (R=–0.558, p=0.016) in IAV patients (Figure 3H). During convalescence, CD1c+ MDC frequencies were increased in blood and lowered in nasopharynx (Figure 3I). Taken together, we showed that the DC compartment also underwent substantial changes in both blood and the nasopharynx during acute IAV infection, potentially suggesting that DCs also received signals to induce migration and/or maturation.

**Figure 3.**
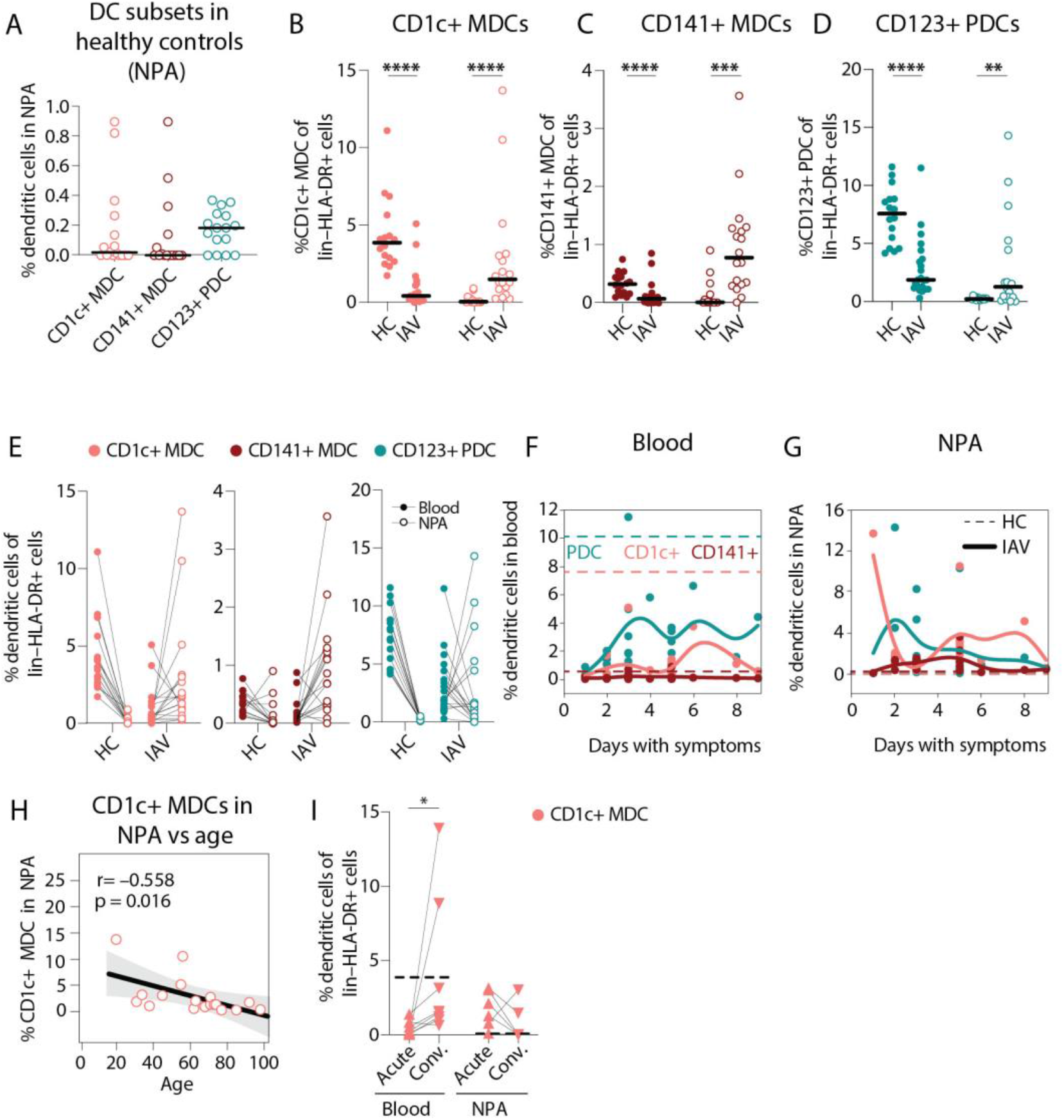
Lower frequencies of dendritic cells in blood but higher frequencies in the nasopharynx in IAV patients compared to healthy controls. (A) Scatter plots shows the frequencies of CD1c+ (coral) and CD141+ (maroon) myeloid DCs (MDCs); and CD123+ PDCs (teal) in the NPA from HCs (n=16). Lines indicate median frequencies. Graphs show frequencies of (B) CD1c+ MDCs, (C) CD141+ MDCs and (D) CD123+ PDCs expressed as a frequency of lin–HLA-DR+ cells in PBMCs and NPA from HCs (n=12) and IAV patients (n=22). Shaded bars represent median values. Differences between IAV patients and HCs were assessed using Mann-Whitney test and considered significant at p<0.05 (**p<0.01, ***p<0.001, ****p<0.0001). (E) displays frequencies of DC subsets from B-D again, with lines connecting values in blood (filled circles) and NPA (open circles) in the same individual. (F-G) Temporal changes in DC subset frequencies as a function of days with symptoms. Solid lines illustrate the temporal variation in the frequencies of PDCs (teal), CD1c+ MDCs (coral) and CD141+ MDCs (maroon) in (F) blood and (G) NPA of IAV patients as compared to median values observed in HCs (dashed lines). (H) Scatter plot and line of fit display bivariate linear regression analysis between age and frequency of CD1c+ MDCs in blood. The shaded area represents the 95% confidence region for the fitted line. R represents Spearman ρ and differences were considered significant at p<0.05. (I) Graph depicts frequencies of CD1c MDCs (coral) in blood (n=8) and the NPA (n=6) in the acute (upward triangles) and convalescent phase (downward triangles) in IAV patients. Dashed lines depict median frequency values from HCs in blood and NPA. Differences between acute and convalescent phase values were assessed using Wilcoxon matched-pairs signed rank test and considered significant at p<0.05 (*p<0.05).

### Monocytes and DCs recruited to the human nasopharynx during IAV infection are more mature

We next analyzed the maturation status of DCs and monocytes in blood and nasopharynx samples from IAV patients and HCs (Figure 4A-D). Cells from HCs had low and comparable levels of surface HLA-DR in both blood and the nasopharynx (Figure 4A). In contrast, in IAV patients, monocytes and DCs in the nasopharynx expressed higher levels of HLA-DR than those in blood (Figure 4A and C). We also found that in IAV patients, nasopharyngeal CD1c+ MDCs and CD141+ MDCs expressed more CD86 than cells in blood (Figure 4D), while nasopharyngeal PDCs showed significant upregulation of CD83 during IAV infection as compared to blood PDCs (Figure 4E). When we compared maturation of CM and MDCs (i.e., CD86 expression) with viral RNA load (i.e., Ct values- number of cycles required to amplify viral RNA), we observed a significant inverse correlation (R=–0.696, p=0.0019), implying that higher viral RNA loads (low Ct value) were associated with increased maturation of CMs (Figure 4F). We also observed a positive correlation between CD86 expression on nasopharyngeal CMs, and both nasopharyngeal CD1c+ MDCs (R=0.735, p=0.0012) and CD141+ MDCs (R=0.832, p=0.0001) (Figures 4G and H, respectively). This indicates that in patients with greater viral RNA load who also have more mature CMs in the nasopharynx, there is a greater likelihood of finding mature MDCs in the nasopharynx as well.

**Figure 4.**
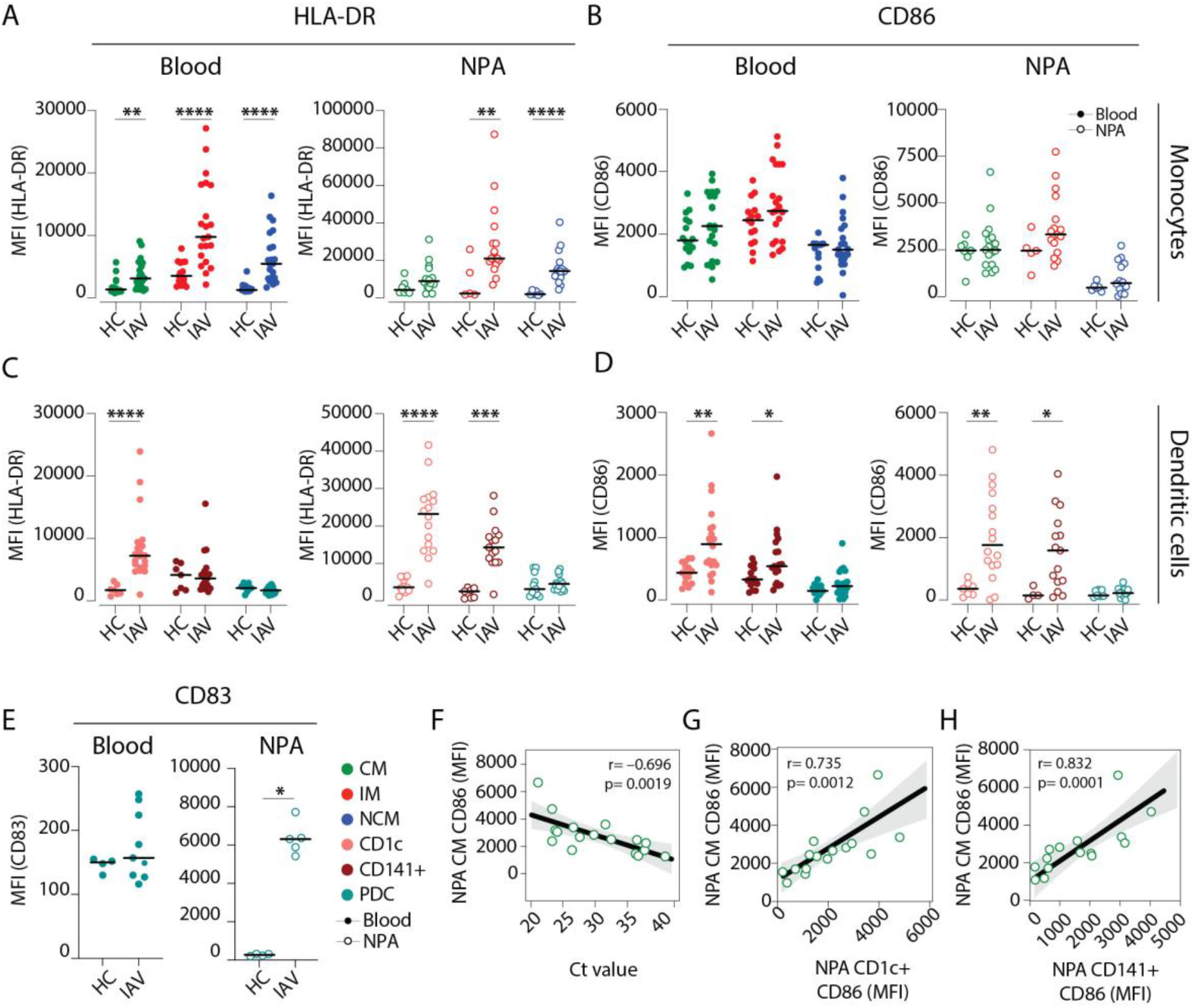
Intermediate monocytes and MDCs in the nasopharynx are more mature in IAV patients than in healthy controls. (**A-D**) Scatter plots depict MFI of (**A, C**) HLA-DR and (**B, D**) CD86 surface expression on (**A-B**) monocyte and (**C-D**) DC subsets in blood (filled circles) and in the NPA (open circles) in HCs (left, n=11) and IAV patients (right, n= 19). (**E**) Scatter plot depicts MFI of CD83 expression on PDCs in blood (filled circles) and in the NPA (open circles). Differences between IAV patients and HCs were assessed by Mann-Whitney test and considered significant at p<0.05. (*p<0.05, **p<0.01, ***p<0.001 and ****p<0.0001). (**F-H**) Scatter plots and lines of fit display bivariate linear regression analysis between monocyte maturation status (CD86 surface expression (MFI)) of NPA CMs in IAV + patients and (**F**) Ct values, (**G**) maturation status (CD86 surface expression (MFI)) of NPA CD1c+ MDCs in IAV patients and (**H**) maturation status (CD86 surface expression (MFI)) of NPA CD141+ MDCs in in IAV patients. The shaded area represents the 95% confidence region for the fitted line. R represents Spearman ρ and differences were considered significant at p<0.05.

### Elevated cytokine levels observed in plasma and the nasopharyngeal secretions correlate with increased monocyte frequencies in respective compartment

Pronounced cytokinemia is a hallmark of severe influenza disease (18). In order to characterize the degree of inflammation in IAV patients, we measured local and systemic cytokine levels. In agreement with earlier reports (2, 3, 29, 30), we observed elevated levels of TNFα, IL-6 and IFNα in nasopharyngeal secretions (Figures 5A-C) as well as in plasma of IAV patients as compared to HCs (Figures 5D-F). We also observed elevated levels of plasma IL-10, IL-15 and IL-18 in IAV patients as compared to HCs (data not shown). We compared soluble TNFα levels against frequencies of monocytes and DCs (i.e., potential cellular sources) at the respective anatomical sites; and found a positive correlation between soluble TNFα and CM frequency, both in blood (Figure 5G); and in the nasopharynx of IAV patients (Figure 5H). However, such correlation was not observed for DCs in blood or NPA (data not shown). Interestingly, we also observed positive associations between age and level of TNFα in plasma and NPA in IAV patients (Figure 5I). Furthermore, we observed that plasma IFNα levels were positively correlated with Ct value (suggesting inverse correlation with viral RNA load) (Figure 5J). Patients with higher viral RNA loads had lower amounts of IFNα in circulation, which may suggest incomplete protection from infection. For a subset of patients, we assessed chemokines (CCL2, CCL3 and CCL7) in circulation and at the site of infection. Interestingly, in blood, elevated plasma levels of CCL2 correlated positively with IM frequencies, which were significantly elevated in IAV patients, indication a role for CCL2 in the changes to the IMs during infection (Figure 5K). Moreover, during convalescence, TNFα and IL-6 were also reduced both in the nasopharynx and in blood suggesting ablation of both local and systemic inflammation (Figures 5L).

**Figure 5.**
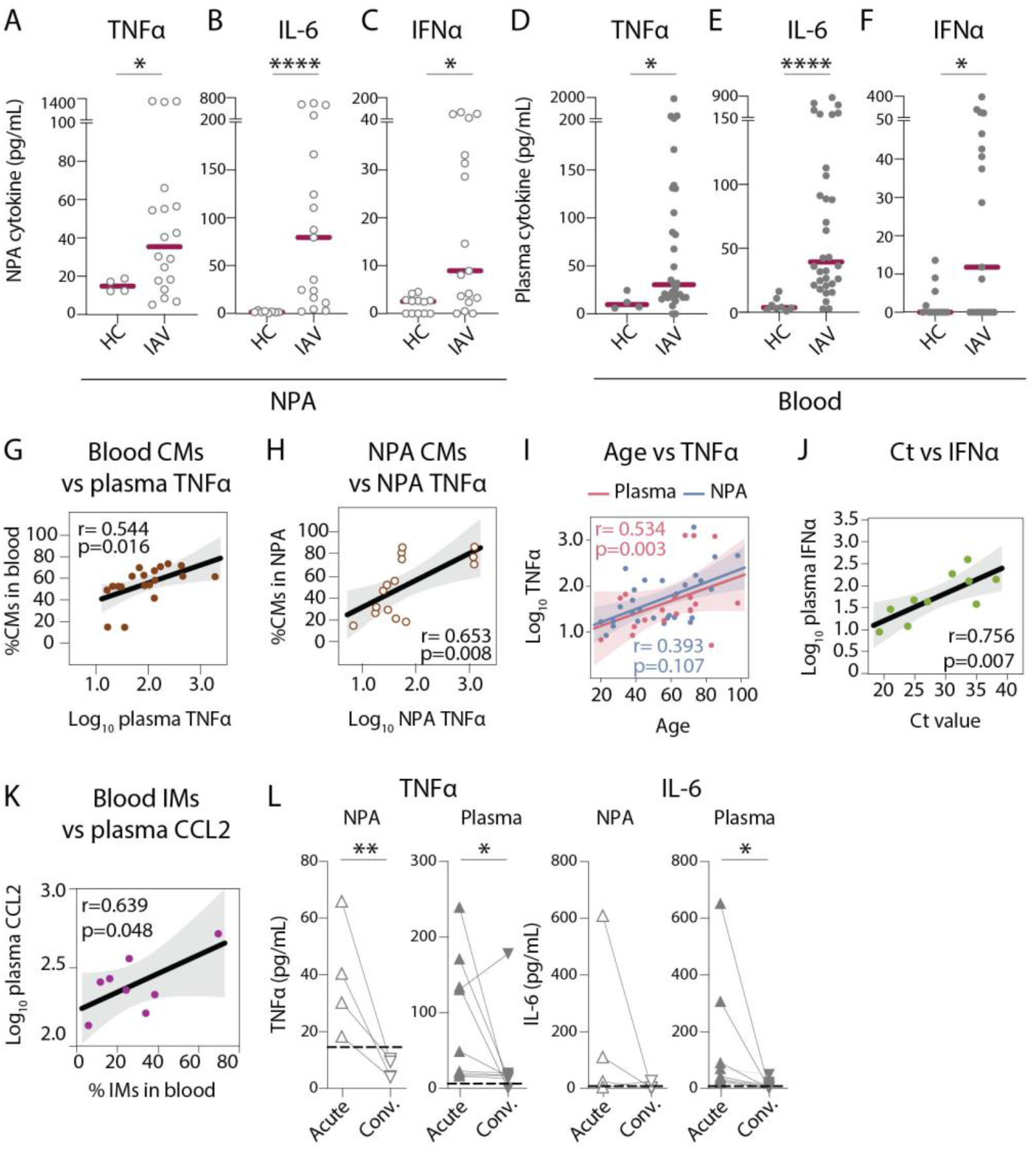
Nasopharyngeal and plasma levels of proinflammatory cytokines TNFα, IL-6 and IFNα are elevated during acute IAV infection. (A-F) Graphs show concentrations of (A and D) TNFα, (B and E) IL-6, (C and F) IFNα in (A-C) NPA (open circles) and (D-F) plasma (filled circles) from HCs (n=12) and IAV patients (n=31) as determined by ELISA. Lines indicate median concentration. Differences between IAV patients and HCs were assessed using Mann-Whitney test and considered significant at p<0.05 (*p<0.05, **p<0.01, ****p<0.0001). (G-K) Scatter plots and lines of fit display bivariate linear regression analysis between variables. The shaded area represents the 95% confidence region for the fitted line. R represents Spearman ρ and differences were considered significant at p<0.05. (G) Log_10_ plasma TNFα values vs CM frequency in blood of IAV patients; (H) log_10_ NPA TNFα vs CM frequency in the NPA of IAV patients; (I) age vs log_10_ TNFα values in plasma (pink) and NPA (blue) in IAV patients; (J) Ct values vs log_10_ IFNα values in IAV patients (light green) and (K) log_10_ plasma CCL2 vs IM frequency in blood of IAV patients. (L) Graphs depicts TNFα and IL-6 levels in the NPA (n=4) and plasma (n=8) during the acute (upward triangles) and convalescent phase (downward triangles) in IAV patients. Dashed lines depict median frequency values from HCs in blood and NPA. Differences between acute and convalescent phase values were assessed using Wilcoxon matched-pairs signed rank test and considered significant at p<0.05 (*p<0.05 and **p<0.01).

### Monocytes are a potent source of systemic TNFα during IAV infection

The localization of mature monocytes and DCs in the nasopharynx during IAV infection, and increased cytokine levels in both compartments led us to question whether the cells in circulation were directly involved in inflammation during ongoing infection, or primarily provided a cache of differentiated cells that can migrate to the site of infection. Limited by the number of viable cells obtained from the nasopharynx, we tested the functional response of circulating monocytes and DCs to in vitro stimulation with a TLR7/8 agonist which mimics ssRNA and then quantified the frequency of TNF-producing cells in each monocyte and DC subset (Figures 6A and B). We observed that blood monocytes and DCs from IAV patients produced TNFα spontaneously, in the absence of any external stimulus, with most of the cytokine coming from monocytes (CMs > IMs > NCMs) (Figure 6C). In contrast, blood monocytes and DCs from HCs only produced TNFα upon stimulation with TLR7/8L (Figure 6D). Importantly, while many of the IAV patients had cells producing TNFα spontaneously, monocytes and DCs remained responsive and had the potential for further increased frequency of TNF-producing cells in response to TLR7/8L stimulation (Figure 6D). In summary, our data suggest that during IAV infection, mature monocytes and DCs accumulate in the nasopharynx, and blood monocytes and DCs function as a general source of TNFα, potentially contributing to the systemic inflammatory effects accompanying influenza infections.

**Figure 6.**
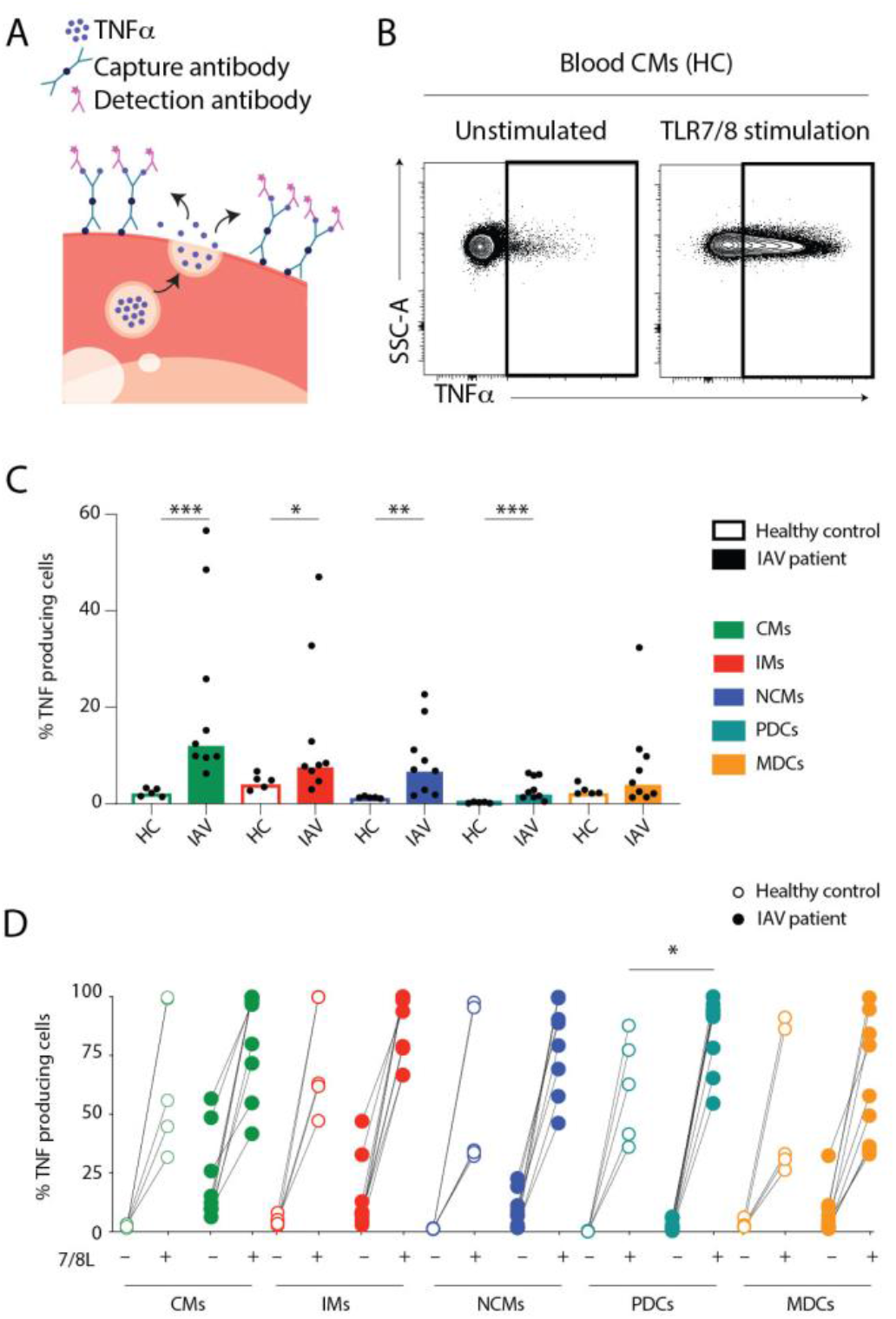
Monocytes and DCs from IAV patients produce TNFα ex vivo without stimulation. (**A**) TNFα release assay: the capture antibody (green) immobilizes secreted TNFα on the cell surface of the TNFα-secreting cell. The PE-labelled detection antibody (pink) together with the phenotypic antibody panel allows detection of TNFα production in individual cell subsets by flow cytometry. (Illustrations were modified from Servier Medical Art, licensed under a Creative Commons Attribution 3.0 Unported License). (**B**) Representative flow cytometry plots of TNFα-producing blood CMs from one HC after 2 hours at 37°C without (unstimulated) or with TLR7/8L stimulation. (**C**) Bar graphs display median frequency of TNFα-producing cells in CMs (green), IMs (red), NCMs (blue), PDCs (teal) and total MDCs (orange) in blood in HCs (open, n=5) and IAV patients (filled, n=9) in the absence of TLR stimulation. Each dot represents an individual donor. (**D**) Graph displays frequency of TNFα-producing cells in monocyte and DC subsets from HCs (open, n=5) and IAV patients (filled n=9) in the absence (–) or presence (+) of TLR7/8L stimulation. Differences between IAV patients and HCs in (**C**) were assessed by Mann-Whitney test and in (**D**) with two-way ANOVA analysis using Sidak’s multiple comparisons test and considered significant at p<0.05. (*p<0.05, **p<0.01 and ***p<0.001).

### Nasopharyngeal aspirates allow assessment of in situ immune responses to mild infections with other respiratory viruses including SARS-CoV-2

In order to determine whether our findings were pathogen-specific to IAV or a reflection of immune responses to respiratory viral infections in general, we analyzed samples from patients with confirmed IBV, RSV or SARS-CoV-2 infections. IBV and RSV patients were closer to IAV patients in age (mean: 53 and 61 years, respectively) and days with symptoms (mean: 5.0 and 6.5 days respectively). In contrast, patients with mild SARS-CoV-2 were younger than IAV patients (mean: 44 years and 59 years respectively) and sought medical attention later: mean 13.2 days with symptoms (SARS-CoV-2) as compared to 3.3 days (IAV) (Table 2). Similar to IAV patients, patients with SARS-CoV-2 and RSV had a higher yield of cells in the nasopharynx than HCs (Figure 7A). The expansion in monocyte and DC frequencies in blood we saw in IAV patients, was not observed in other infections. Interestingly, in the nasopharynx, monocytes and DCs were significantly elevated during IAV infection, but not during IBV infection (Figure 7B). Within the monocyte compartment, we noted differences between the different pathogens. IMs were not increased during IBV or SARS-CoV-2 infection but were increased during RSV infection in both blood and the nasopharynx (Figure 7C). The DC compartment too showed differences between the groups in NPA. In blood, all DC subsets were decreased in all groups (Figure 7D). CD1c+ MDCs were increased in the nasopharynx only during IAV and SARS-CoV-2 infections. Interestingly, CD141+ MDCs were significantly increased in the nasopharynx only during IAV infection. PDCs in NPA were increased in all groups.

**Figure 7.**
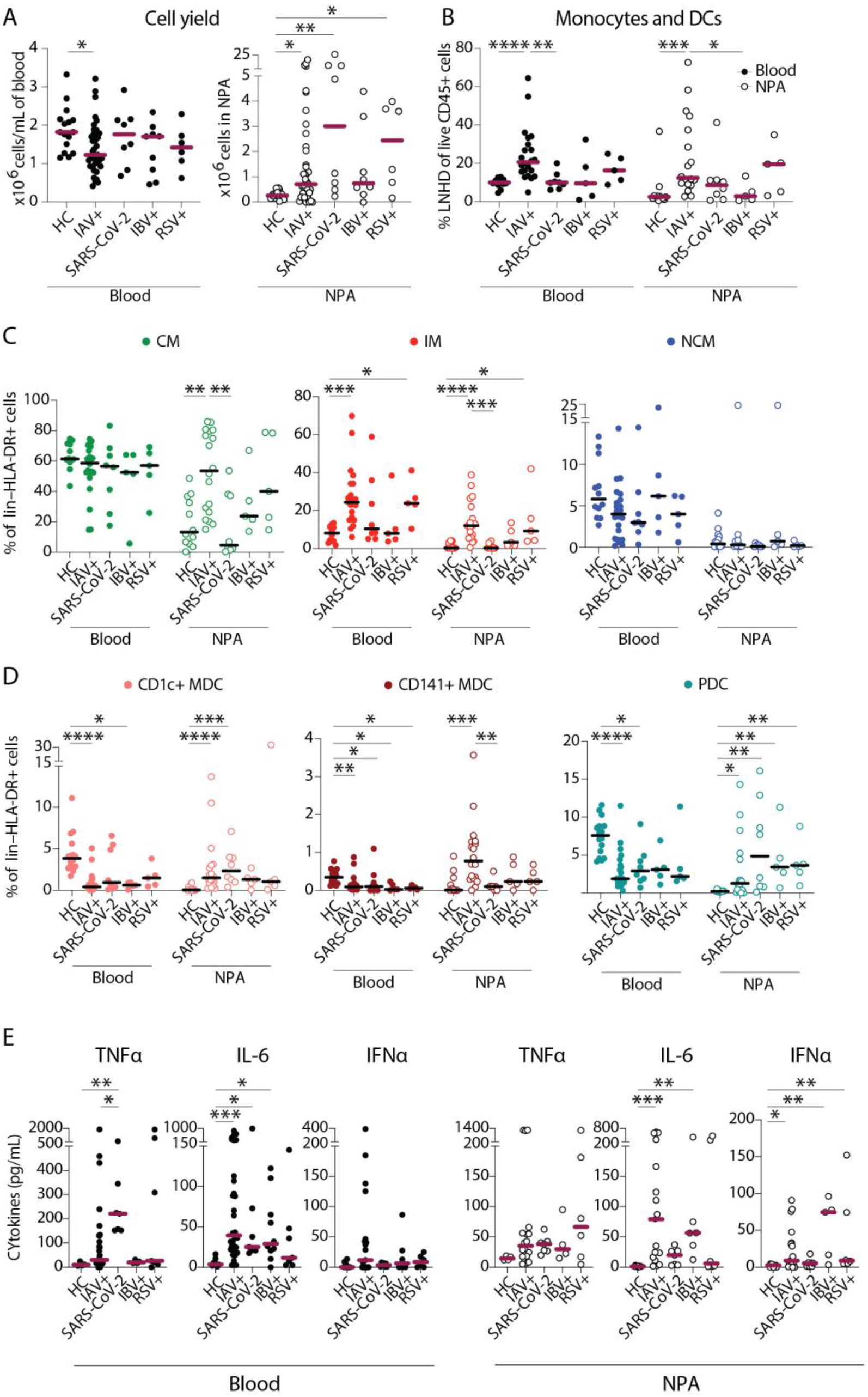
SARS-CoV-2 infection elicits a distinct pattern of innate immune response as compared to IAV infection. (**A-B**) Scatter plots show data from individual subjects and lines indicate median values. (**A**) Cell yield per mL blood (x10^6^ PBMCs, filled circles) and total NPA cells (x10^6^ cells, open circles) obtained from patients and HCs. (**B**) Frequency of lin–HLA-DR+cells of live CD45+ cells in blood (filled circles) and NPA (open circles) from HCs and patients. (**C**-**D**) Scatter plots show frequencies of (**D**) monocyte subsets (CM: green, IM: red, NCM: blue) and (**E**) DC subsets (CD1c+: coral, CD141: maroon, PDC: teal) in PBMCs (filled circles) and NPA (open circles) from HCs and patients. Graphs show data from individual subjects and lines represent median values. Differences between HCs or IAV patients and other patient groups were assessed using Kruskal-Wallis test with Dunn’s multiple comparisons test and considered significant at p<0.05 (*p<0.05, **p<0.01, ***p<0.001 and ****p<0.0001). (**E**) Graphs show concentrations of TNFα, IL-6 and IFNα in plasma (filled circles) and NPA (open circles) from HCs and patients as determined by ELISA. Lines indicate median concentration. Differences between IAV patients and HCs were assessed using Mann-Whitney test and considered significant at p<0.05 (*p<0.05, **p<0.01 and ***p<0.001).

In plasma, individuals with SARS-CoV-2 infection had elevated levels of TNFα in comparison with HCs and IAV patients (Figure 7E). SARS-CoV-2 infection was associated with elevated levels of IL-6 compared to HCs, but not compared to the IAV and IBV groups. In NPA, only the IAV and IBV groups had increased levels of IL-6. Despite having increased frequencies of PDCs in NPA, nasopharyngeal levels of IFNα were not elevated in SARS-CoV-2 patients. Together, these data show different patterns of monocyte and DC engagement in the nasopharynx and in blood, and also between IAV, IBV, RSV and SARS-CoV-2, suggesting a requirement for further scrutiny.

## DISCUSSION

In this study, we mapped monocyte and DC distribution and function in blood and in the nasopharynx (the initial site of infection) of patients with ongoing IAV, IBV, RSV or SARS-CoV-2 infections and healthy controls. Several studies have examined immune responses during severe flu, in particular, focusing on patients sampled during or immediately following the 2009 H1N1 pandemic (2, 3, 31, 40), or patients hospitalized with severe respiratory symptoms (2, 29–31, 40); and more recently, the immune dysregulation during severe COVID-19 (41–44). Here, we elucidated the innate myeloid cell composition and responses in a cohort with relatively mild symptoms, advanced age and underlying comorbidities, typical of seasonal influenza. Finally, we showed that our methods can be adapted to study immune responses in other viral infections, including the current COVID-19 pandemic.

Flow cytometric characterization revealed an influx of monocytes and DCs to the nasopharynx during infection, in line with previous reports in more severe IAV patients (3, 29, 30, 45). Interestingly, age appeared to differentially skew the IM response in IAV patients. Despite having higher baseline frequencies of circulating CD16-expressing monocytes (46), older patients have fewer IMs in blood during acute IAV infection and concurrently more IMs in the nasopharynx. The increase in IMs suggests a more inflammatory milieu at the site of infection in older patients, perhaps contributing to disease severity as previously suggested in pandemic influenza (2–5). Despite the general increase in IM frequencies, CMs remained the most frequent monocyte/DC subset in either compartment implying a functional role for CMs during IAV infection. Similar to studies in paediatric patients (29, 30), we also found that over the course of the illness, DCs appeared to migrate from blood to infiltrate the nasopharynx. However, older patients displayed a weaker recruitment of CD1c+ MDCs. In vitro studies have shown that monocytes can differentiate into type I IFN producing monocyte-derived DCs in response to IAV exposure (47). It is therefore possible that CMs recruited to the nasopharynx in older individuals preferentially differentiate into IMs rather than DCs. Moreover, CD1c+ DC function is critical for clearance of IAV infection (35). Therefore, diminished recruitment of CD1c+ MDCs and or reduced in situ DC differentiation (and therefore delayed or attenuated CD8+ T cell responses) may also contribute more severe disease in older patients.

During acute disease, monocytes present in/recruited to the nasopharynx likely contribute to sustained DC recruitment by secreting TNFα, CCL2, CCL3 and CCL7 locally (3, 29, 40). Due to the limited availability of cells in the NPA samples, demonstrating cytokine secretion from nasopharyngeal cells was not possible. However, we noted that in the nasopharynx, CMs, CD1c+ MDCs and CD141+ MDCs all appeared to mature by upregulating HLA-DR and/or CD86 expression as compared to HCs, suggesting local activation, likely in response to the inflammatory milieu and/or virus exposure. CD86, a critical costimulatory molecule and ligand for CD28 and CTLA-4, is upregulated on monocytes and DCs upon virus infection in vitro and in vivo (48). Lower expression of CD86 on circulating myeloid cells as compared to nasopharyngeal cells during IAV infection suggests migration of mature cells to the nasopharynx, and/or *in situ* differentiation of monocytes to induced-DCs (that may not upregulate maturation markers) in blood during acute IAV infection as previously speculated (47). A non-human primate model of chronic simian immunodeficiency virus (SIV) infection revealed sustained mobilization of MDCs from the bone marrow via blood to the intestinal mucosa, where MDCs remained activated and eventually underwent apoptosis (49). In IAV infection, monocytes and DCs are likely recruited to the nasopharynx for a shorter period of time, where they face a similar fate. Some DCs likely traffic antigens to the lymph node to support adaptive responses (21, 35).

Nasopharyngeal CMs were also more mature in patients with higher viral RNA loads and their recruitment and/or differentiation may be aided by local cytokine production (TNFα and IL-6). Key innate inflammatory cytokines, including TNFα, IL-6, IFNα, IL-10, IL-15, and IL-18, were all significantly elevated early during infection (days 1-5) compared to healthy controls. TNFα levels strongly correlated with the presence of CMs in both blood and nasopharynx; despite the significant expansion of IMs in blood, supporting previous studies (2, 14, 31, 50). We also demonstrated that be circulating CMs was the most frequent cellular source of TNFα among blood monocytes and DCs, while the TNFα at the site of infection may come from both CMs and the IMs (both subsets found in increased frequencies). *Ex vivo*, at steady state, human blood CMs and IMs have comparable TNFα secretion irrespective of the nature of stimulation (14). IAV infection, therefore, may skew monocyte subset function, in addition to monocyte distribution and maturation. The broad range of TNFα, IL-6 and IFNα responses seem among the IAV patients suggest that seasonal IAV strains are associated with milder cytokine responses than those observed during infections with 2009 H1N1 pandemic influenza or highly pathogenic avian H5N1 strains (3, 10, 31, 51). Within the setting of seasonal influenza however, older patients had more IMs in the NPA and displayed stronger TNFα responses, locally and systemically.

During IAV infection, the significant expansion of IMs in blood and the nasopharynx could be mediated by the presence of CCL2, a chemokine critical for monocyte chemotaxis (52). Upon CCL2 ligation, CCR2 facilitates CM and IM adhesion and transmigration from blood to tissues (53). Therefore, in IAV patients, CCL2 may contribute to recruitment of monocytes and DCs from circulation to the nasopharynx to aid inflammation (3, 50). Furthermore, elevated IFNα in circulation correlated with reduced viral RNA load, although we saw no evidence of this locally at the site of infection; suggesting that perhaps lower systemic IFN responses can indicate impaired control of viral replication in the nasopharynx, as previously reported (31). This finding also supports previous studies showing that attenuated RIG-I signalling impairs IFN responses in the elderly (54) and very young (55), both groups at high risk of influenza-associated mortality.

During convalescence, monocyte and DC frequencies, and cytokine levels normalized in blood and the nasopharynx to levels seen in healthy individuals. The local and systemic inflammation observed, therefore, appears limited to the acute phase of disease and wanes during convalescence. Unlike patients infected with the pandemic 2009 H1N1 IAV strain, where long lasting immune perturbations were documented (28); the repercussions of relatively mild seasonal IAV infections appear short-lived. This is also in contrast to in vivo studies which demonstrated prolonged DC activation and disruption of DC subset reconstitution in the lungs following IAV infection (56).

In April-May 2020, during the COVID-19 pandemic, the approach and methods described here allowed us to implement this study plan in a rapidly evolving pandemic. Although the patient cohorts were not identical, this endeavour proved the feasibility of using nasopharyngeal aspiration to assess local immune responses during respiratory viral infections. We observed that patients with IBV, RSV or SARS-CoV-2 also displayed an influx of cells to the nasopharynx during infection. Despite annual reoccurrence of IBV and RSV infections, detailed investigations of the innate cells at the site of infection in patients are quite rare (29, 30), and the technique we describe allows for minimally invasive longitudinal measurement of cellular and molecular aspects of infection. Patients with mild SARS-CoV-2 infection have not been studied as extensively as patients with severe or fatal COVID-19. The cell influx to the nasopharynx in COVID-19 patients did not appear to be due to monocytes but rather due to CD1c+ MDCs and PDCs, which were reduced in blood, in line with previous reports (41, 57). Also in line with previous studies (42–44), we found elevated levels of IL-6 in plasma, and also of TNFα. As different SARS-CoV-2 variants emerge, with concurrently increasing global vaccination rates, incomplete neutralization and/or protection and herd immunity may result in recurring, mild SARS-CoV- 2 infections. Understanding site-of-infection responses to mild SARS-CoV-2 infections is therefore increasingly relevant, and can be accomplished by the methods we describe in our study.

The present study augments our current understanding of the role of monocyte and DC subsets during human respiratory viral infections by highlighting unique dynamics and location-specific functions for each subset over the course of infection. Acute human IAV infection is characterized by an expansion of IMs and monocyte-mediated cytokinemia driving monocyte and DC migration from blood to the nasopharynx, the initial site of IAV infection. Older patients with comorbidities display a recruitment pattern skewed towards IMs and increased TNF rather than CD1c+ MDCs which may contribute to more severe disease and longer duration of hospitalization observed in these patients. IBV, RSV and mild SARS-CoV-2 infections elicit different patterns on immune cell recruitment to the nasopharynx as compared to IAV infections. We also illustrate the value of comparative high-resolution studies of immune cells in blood and at the site of infection, in order to fully understand their individual contributions to disease and how they orchestrate inflammation synergistically. Similar studies, carried out longitudinally across tissues, will aid resolution of these findings, and allow multivariate modelling of biomarkers of disease severity. Therapeutic approaches which allow selective modulation of monocyte and DC redistribution, maturation and cytokine/chemokine function may hold the key to reducing influenza-associated disease burden and mortality in the future.

## METHODS

### Study subjects

Patients seeking medical attention for influenza-like illness at the Emergency Department at the Karolinska University Hospital in Stockholm, Sweden during three consecutive influenza seasons (January - March) of 2016-2018 were recruited to the study following informed consent. The inclusion criteria for enrolment of patients were (1) age >18 years, (2) no known immunodeficiency, (3) not taking antibiotics, immunomodulatory or anti-inflammatory medication at time of inclusion, (4) presenting with fever and at least one of the following symptoms of influenza-like illness: cough, nasal congestion, headache or muscle ache. Convalescent samples were collected after at least 4 weeks, ensuring absence of respiratory symptoms in the prior week. Healthy volunteers were recruited and sampled similarly outside of influenza season. During the COVID-19 pandemic, additional patients were included between April - May 2020 at the Infectious Diseases ward at the Karolinska University Hospital or the Haga Outpatient Clinic (Haga Närakut) in Stockholm, Sweden, as well as mild/asymptomatic household contacts of patients with confirmed COVID-19 were screened by PCR and enrolled if positive. Clinical data were obtained from the patients and medical records (Table 1) and are extensively discussed in a previous publication (58). Total burden of comorbidities was assessed using the CCI (59).

The severity of disease was categorized using the respiratory domain of the sequential organ failure assessment score (SOFA) (60). In the absence of arterial partial pressure of oxygen (PaO2), peripheral transcutaneous haemoglobin saturation (SpO2) was used instead to calculate a modified SOFA score (mSOFA) (61). Fraction of inspired oxygen (FiO_2_) estimation based on O_2_ flow was done in accordance with the Swedish Intensive Care register definition (62). Mild disease was defined as PaO_2_/FiO_2_ (PFI) >53 kPa (>400 mmHg) or SpO_2_/FiO_2_ (SFI) >400. Moderate disease was defined as PFI >27-53 kPa (>200-400 mmHg) or SFI 235-400. The disease severity score is more extensively described in a previous publication (38).

### Sample collection

Blood, nasal swabs and nasopharyngeal aspirates (NPA) were obtained from all patients (acute and convalescent phase samples) and healthy controls (Figure 1A). Briefly, up to 30mL venous blood was collected in Vacutainer® tubes containing EDTA, for blood counts and PBMC isolation. Nasopharyngeal swabs (Sigma Virocult®) were collected for diagnostic qPCR. NPA samples were collected into a vacuum trap by inserting a thin catheter through the naris, deep into the nasopharynx and applying gentle suction for 1-3 minutes. The vacuum trap and tubing were rinsed out with 3mL sterile PBS. All samples were processed within 2 hours of sampling.

### Diagnostic tests to determine etiology of infection

Nasal swab samples were analyzed for influenza A virus (IAV), influenza B virus (IBV) and respiratory syncytial virus (RSV) by real-time PCR using the commercial Simplexa system^TM^ (63), as well as bacteria (by culture methods). The tests were performed at the Department of Clinical Microbiology, Karolinska University Laboratory as part of routine diagnostics for respiratory viral infections. Cycle threshold (Ct) values from the Simplexa^TM^ assay were considered as semi-quantitative measures of virus levels in statistical analyses. Bacterial culture results were retrieved wherever available from the Department of Clinical Microbiology at the Karolinska University Hospital. Convalescent individuals and healthy controls were confirmed IAV– by qPCR. SARS-CoV-2 infection was diagnosed similarly using the GeneXpert SARS-CoV-2 detection system (Cepheid). Supplementary data on bacterial cultures were retrieved from the microbiology lab/clinical records.

### Isolation of cells from blood and nasopharyngeal aspirates

Blood samples were centrifuged at 800g/10 min/room temperature (RT) and plasma was frozen at – 20°C. The blood volume was reconstituted with sterile PBS and PBMCs were obtained by density-gradient centrifugation using Ficoll-Paque Plus (GE Healthcare) after centrifugation at 900g/25 min/RT (without brake). NPA samples were centrifuged at 400g/5 min/RT and the supernatant was frozen at – 20°C. The cells were washed with 5mL sterile PBS to remove mucus, by filtering through a 70μm cell strainer followed by centrifugation for 400g/5 min/RT. Cell counts and viability were assessed using Trypan Blue (Sigma) exclusion and an automated Countess cell counter (Invitrogen) or manual counting using a light microscope (when the counter was unable to distinguish live and dead NPA cells). Cells were used fresh for flow cytometry (all NPA cells when samples contained >10^5^ cells and had ≥70% viability; and at least 1x10^6^ PBMCs) or frozen in RNALater (<10^5^ NPA cells) (ThermoFisher). Excess PBMCs were cryopreserved in FBS (Gibco) + 10% DMSO (Sigma) and stored in liquid nitrogen.

### Flow cytometry analysis

Phenotypic analysis was performed on NPA cells and PBMCs using Live/Dead Blue; lineage markers CD3 (SK7; BD), CD19 (HIB19; Biolegend), CD20 (L27, BD), CD45 (HI30; BD), CD56 (HCD56, BD) and CD66abce (TET2, Miltenyi Biotec); HLA-DR (TU36, Life technologies), CD14 (M5E2, BD), CD16 (3GE, Biolegend), CD11c (B-Ly6, BD), CD1c (AD5-8E7; Miltenyi), CD141 (AD5-14H12; Miltenyi), CD123 (7G3; BD); maturation markers CD83 (HB15e, Biolegend) and CD86 (2331; BD); adhesion marker CD62L (SK11, BD); and migration markers CCR2 (K036C2, Biolegend) and CCR7 (150503: BD); and fixed with 1% paraformaldehyde for flow cytometry. Samples were acquired on an LSRFortessa flow cytometer (BD Biosciences) and analyzed using FlowJo software v10 (Tree Star).

### TNF-α release assay

TNFα secretion from fresh PBMCs in response to TLR stimulation was assessed using the TNF-α Secretion Assay-Detection Kit (PE) (130-091-268, Miltenyi Biotec) according to the manufacturer’s instructions. TNFα secretion was measured over 2 hours of incubation at 37°C with shaking (200 rpm) in the presence or absence of 1 μg/mL 3M-019 (7/8L) (Invivogen). Briefly, the capture antibody immobilized all secreted TNFα on the surface of the cell, and the detection antibody (PE-labelled) was incorporated into the above panel of antibodies to determine the cellular source of TNFα. Secretion of TNFα was quantified using flow cytometry.

### ELISA and Luminex

Plasma samples and NPA supernatants were assayed for soluble markers using ELISA and Luminex. Human IFNα All Subtype ELISA was performed according to the manufacturer’s instructions (PBL Assay Science). TNFα and IL-6 ELISAs were performed using DuoSet® kits (R&D Systems). Luminex assays were performed using custom-designed 9-plex (IFNγ, IL-8, IL-10, IL-18, CCL2, CCL3, CCL7, IL-1β and IL-12p70) kit (R&D Systems) and analyzed on the Bio-Plex® 200 instrument (Bio-Rad).

### Statistics

Data were analyzed using GraphPad Prism version 6.0 (GraphPad Software) and JMP®, version 14.2. (SAS Institute Inc., Cary, NC, 1989-2019). Differences between frequencies in IAV patients and HCs were assessed using nonparametric tests after assessing normality, using the Mann-Whitney test (at 95% confidence intervals). For comparisons between exposure conditions, two-way ANOVA with Sidak’s multiple comparisons test was applied. For comparisons between acute and convalescent phase data, Wilcoxon matched-pairs signed rank test was used. Bivariate and multivariate linear regression analysis was performed using JMP®, choosing Spearman’s rank correlation coefficient for nonparametric analyses. Differences between HCs or IAV patients and other patient groups were assessed using nonparametric tests after assessing normality, using the Kruskal-Wallis test with Dunn’s multiple comparisons test (at 95% confidence intervals). Data were considered significant at p<0.05.

### Study approval

Informed consent was obtained from all patients and volunteers following verbal and written information. The study was approved by the Swedish Ethical Review Authority (No. 2015/1949-31/4) and performed according to the Declaration of Helsinki.

## ADDITIONAL SUPPLEMENTARY MATERIAL

**Supplementary Table 1.**
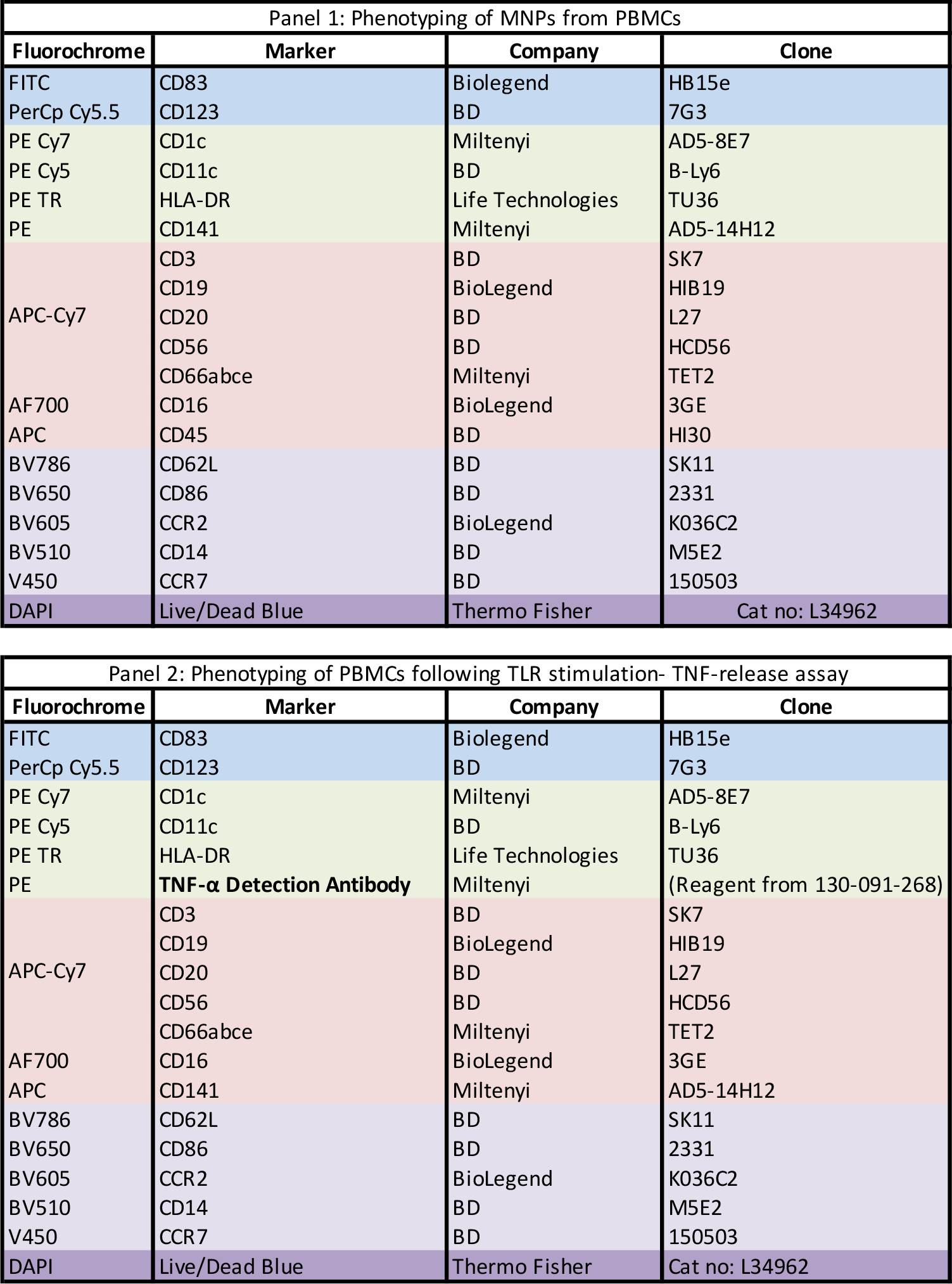
**Flow cytometry panels.**

## Data Availability

All data produced in the present study are available upon reasonable request to the authors.

## ACKNOWLEDGMENTS

We thank the patients and healthy volunteers who have contributed clinical material to this study. We would like to thank Adeline Mawa, Kevin Fallahi, Marcus Nordin, Roosa Vaitiniemi, Eric Åhlberg and the research nurses at the Emergency and Infectious Diseases Departments for technical assistance. This work was supported by grants to AS-S from the Swedish Research Council (VR), the Swedish Heart- Lung Foundation, the Bill and Melinda Gates Foundation, the Swedish Childhood Cancer Fund and Karolinska Institutet.

## AUTHOR CONTRIBUTIONS

SV, NJ, AF and AS-S planned the study. SV, MY, SF-J and SL performed experiments. SF-J, BÖ, KS, AF and NJ included and sampled patients, and collected clinical data. JA provided relevant pseudonymized patient clinical data. SV, MY, SF-J, BÖ, SL, MA and AS-S analyzed data. SV and AS- S prepared figures and wrote the manuscript. All co-authors edited the manuscript.

## REFERENCES

1. WHO. Factsheet: Influenza (Seasonal). 2018.

2. Cole SL, Dunning J, Kok WL, Benam KH, Benlahrech A, Repapi E, et al. M1-like monocytes are a major immunological determinant of severity in previously healthy adults with life-threatening influenza. JCI Insight. 2017;2(7):e91868.

3. Oshansky CM, Gartland AJ, Wong SS, Jeevan T, Wang D, Roddam PL, et al. Mucosal immune responses predict clinical outcomes during influenza infection independently of age and viral load. Am J Respir Crit Care Med. 2014;189(4):449–62.

4. Kuiken T, Taubenberger JK. Pathology of human influenza revisited. Vaccine. 2008;26:D59–D66.

5. Kuiken T, van den Brand J, van Riel D, Pantin-Jackwood M, Swayne DE. Comparative pathology of select agent influenza a virus infections. Vet Pathol. 2010;47(5):893–914.

6. Zhou F, Yu T, Du R, Fan G, Liu Y, Liu Z, et al. Clinical course and risk factors for mortality of adult inpatients with COVID-19 in Wuhan, China: a retrospective cohort study. The Lancet. 2020;395(10229):1054–62.

7. Tellier R. Review of Aerosol Transmission of Influenza A Virus. Emerg Infect Dis. 2006;12(11):1657–62.

8. Sanders CJ, Doherty PC, Thomas PG. Respiratory epithelial cells in innate immunity to influenza virus infection. Cell Tissue Res. 2011;343(1):13–21.

9. Stramer SL, Collins C, Nugent T, Wang X, Fuschino M, Heitman JW, et al. Sensitive detection assays for influenza RNA do not reveal viremia in US blood donors. J Infect Dis. 2012;205(6):886–94.

10. Sakabe S, Iwatsuki-Horimoto K, Takano R, Nidom CA, Le M, Nagamura-Inoue T, et al. Cytokine production by primary human macrophages infected with highly pathogenic H5N1 or pandemic H1N1 2009 influenza viruses. J Gen Virol. 2011;92(Pt 6):1428–34.

11. Wang J, Nikrad MP, Travanty EA, Zhou B, Phang T, Gao B, et al. Innate immune response of human alveolar macrophages during influenza A infection. PLoS One. 2012;7(3):e29879.

12. Kwissa M, Nakaya HI, Onlamoon N, Wrammert J, Villinger F, Perng GC, et al. Dengue virus infection induces expansion of a CD14(+)CD16(+) monocyte population that stimulates plasmablast differentiation. Cell Host Microbe. 2014;16(1):115–27.

13. Sporri R, Reis e Sousa C. Inflammatory mediators are insufficient for full dendritic cell activation and promote expansion of CD4+ T cell populations lacking helper function. Nat Immunol. 2005;6(2):163–70.

14. Boyette LB, Macedo C, Hadi K, Elinoff BD, Walters JT, Ramaswami B, et al. Phenotype, function, and differentiation potential of human monocyte subsets. PLoS One. 2017;12(4):e0176460.

15. Jakubzick C, Gautier EL, Gibbings SL, Sojka DK, Schlitzer A, Johnson TE, et al. Minimal differentiation of classical monocytes as they survey steady-state tissues and transport antigen to lymph nodes. Immunity. 2013;39(3):599–610.

16. Patel AA, Zhang Y, Fullerton JN, Boelen L, Rongvaux A, Maini AA, et al. The fate and lifespan of human monocyte subsets in steady state and systemic inflammation. J Exp Med. 2017;214(7):1913–23.

17. Fingerle G, Pforte A, Passlick B, Blumenstein M, Strobel M, Ziegler-Heitbrock L. The novel subset of CD14+:CD16+ blood monocytes is expanded in sepsis patients. Blood. 1993;82(10):3170–6.

18. Tisoncik JR, Korth MJ, Simmons CP, Farrar J, Martin TR, Katze MG. Into the Eye of the Cytokine Storm. Microbiol Mol Biol R. 2012;76(1):16–32.

19. Hemont C, Neel A, Heslan M, Braudeau C, Josien R. Human blood mDC subsets exhibit distinct TLR repertoire and responsiveness. J Leukoc Biol. 2013;93(4):599–609.

20. Jongbloed SL, Kassianos AJ, McDonald KJ, Clark GJ, Ju X, Angel CE, et al. Human CD141+ (BDCA-3)+ dendritic cells (DCs) represent a unique myeloid DC subset that cross-presents necrotic cell antigens. J Exp Med. 2010;207(6):1247–60.

21. Cella M, Jarrossay D, Facchetti F, Alebardi O, Nakajima H, Lanzavecchia A, et al. Plasmacytoid monocytes migrate to inflamed lymph nodes and produce large amounts of type I interferon. Nat Med. 1999;5(8):919–23.

22. Baharom F, Thomas S, Rankin G, Lepzien R, Pourazar J, Behndig AF, et al. Dendritic Cells and Monocytes with Distinct Inflammatory Responses Reside in Lung Mucosa of Healthy Humans. J Immunol. 2016;196(11):4498–509.

23. Alcantara-Hernandez M, Leylek R, Wagar LE, Engleman EG, Keler T, Marinkovich MP, et al. High- Dimensional Phenotypic Mapping of Human Dendritic Cells Reveals Interindividual Variation and Tissue Specialization. Immunity. 2017;47(6):1037–50 e6.

24. Smed-Sorensen A, Chalouni C, Chatterjee B, Cohn L, Blattmann P, Nakamura N, et al. Influenza A virus infection of human primary dendritic cells impairs their ability to cross-present antigen to CD8 T cells. PLoS Pathog. 2012;8(3):e1002572.

25. Diao H, Cui G, Wei Y, Chen J, Zuo J, Cao H, et al. Severe H7N9 infection is associated with decreased antigen-presenting capacity of CD14+ cells. PLoS One. 2014;9(3):e92823.

26. Baharom F, Thomas S, Bieder A, Hellmer M, Volz J, Sandgren KJ, et al. Protection of human myeloid dendritic cell subsets against influenza A virus infection is differentially regulated upon TLR stimulation. J Immunol. 2015;194(9):4422–30.

27. Jochems SP, Marcon F, Carniel BF, Holloway M, Mitsi E, Smith E, et al. Inflammation induced by influenza virus impairs human innate immune control of pneumococcus. Nat Immunol. 2018;19(12):1299–308.

28. Lichtner M, Mastroianni CM, Rossi R, Russo G, Belvisi V, Marocco R, et al. Severe and persistent depletion of circulating plasmacytoid dendritic cells in patients with 2009 pandemic H1N1 infection. PLoS One. 2011;6(5):e19872.

29. Gill MA, Long K, Kwon T, Muniz L, Mejias A, Connolly J, et al. Differential recruitment of dendritic cells and monocytes to respiratory mucosal sites in children with influenza virus or respiratory syncytial virus infection. J Infect Dis. 2008;198(11):1667–76.

30. Gill MA, Palucka KA, Barton T, Ghaffar F, Jafri H, Banchereau J, et al. Mobilization of Plasmacytoid and Myeloid Dendritic Cells to Mucosal Sites in Children with Respiratory Syncytial Virus and Other Viral Respiratory Infections. J Infect Dis. 2005;2005(191):1105–15.

31. Dunning J, Blankley S, Hoang LT, Cox M, Graham CM, James PL, et al. Progression of whole-blood transcriptional signatures from interferon-induced to neutrophil-associated patterns in severe influenza. Nat Immunol. 2018;19(6):625–35.

32. Vangeti S, Gertow J, Yu M, Liu S, Baharom F, Scholz S, et al. Human Blood and Tonsil Plasmacytoid Dendritic Cells Display Similar Gene Expression Profiles but Exhibit Differential Type I IFN Responses to Influenza A Virus Infection. J Immunol. 2019;202(7):2069–81.

33. McGill J, Van Rooijen N, Legge KL. Protective influenza-specific CD8 T cell responses require interactions with dendritic cells in the lungs. J Exp Med. 2008;205(7):1635–46.

34. Mount AM, Belz GT. Mouse Models of Viral Infection: Influenza Infection in the Lung. In: Naik S, editor. Dendritic Cell Protocols Methods in Molecular Biology (Methods and Protocols). 595: Humana Press. ; 2010. p. 299–318.

35. GeurtsvanKessel CH, Willart MA, van Rijt LS, Muskens F, Kool M, Baas C, et al. Clearance of influenza virus from the lung depends on migratory langerin+CD11b- but not plasmacytoid dendritic cells. J Exp Med. 2008;205(7):1621–34.

36. Segura E, Durand M, Amigorena S. Similar antigen cross-presentation capacity and phagocytic functions in all freshly isolated human lymphoid organ-resident dendritic cells. J Exp Med. 2013;210(5):1035–47.

37. Mick E, Kamm J, Pisco AO, Ratnasiri K, Babik JM, Castaneda G, et al. Upper airway gene expression reveals suppressed immune responses to SARS-CoV-2 compared with other respiratory viruses. Nat Commun. 2020;11(1):5854.

38. Falck-Jones S, Vangeti S, Yu M, Falck-Jones R, Cagigi A, Badolati I, et al. Functional monocytic myeloid-derived suppressor cells increase in blood but not airways and predict COVID-19 severity. J Clin Invest. 2021;131(6).

39. Cagigi A, Yu M, Österberg B, Svensson J, Falck-Jones S, Vangeti S, et al. Airway antibodies emerge according to COVID-19 severity and wane rapidly but reappear after SARS-CoV-2 vaccination. JCI Insight. 2021.

40. Marion T, Elbahesh H, Thomas PG, DeVincenzo JP, Webby R, Schughart K. Respiratory Mucosal Proteome Quantification in Human Influenza Infections. PLoS One. 2016;11(4):e0153674.

41. Zingaropoli MA, Nijhawan P, Carraro A, Pasculli P, Zuccala P, Perri V, et al. Increased sCD163 and sCD14 Plasmatic Levels and Depletion of Peripheral Blood Pro-Inflammatory Monocytes, Myeloid and Plasmacytoid Dendritic Cells in Patients With Severe COVID-19 Pneumonia. Front Immunol. 2021;12:627548.

42. Del Valle DM, Kim-Schulze S, Huang HH, Beckmann ND, Nirenberg S, Wang B, et al. An inflammatory cytokine signature predicts COVID-19 severity and survival. Nat Med. 2020;26(10):1636–43.

43. Merad M, Martin JC. Pathological inflammation in patients with COVID-19: a key role for monocytes and macrophages. Nat Rev Immunol. 2020;20(6):355–62.

44. Mudd PA, Crawford JC, Turner JS, Souquette A, Reynolds D, Bender D, et al. Distinct inflammatory profiles distinguish COVID-19 from influenza with limited contributions from cytokine storm. Sci Adv. 2020;6(50).

45. Herold S, von Wulffen W, Steinmueller M, Pleschka S, Kuziel WA, Mack M, et al. Alveolar epithelial cells direct monocyte transepithelial migration upon influenza virus infection: impact of chemokines and adhesion molecules. J Immunol. 2006;177(3):1817–24.

46. Patin E, Hasan M, Bergstedt J, Rouilly V, Libri V, Urrutia A, et al. Natural variation in the parameters of innate immune cells is preferentially driven by genetic factors. Nat Immunol. 2018;19(3):302–14.

47. Cao W, Taylor AK, Biber RE, Davis WG, Kim JH, Reber AJ, et al. Rapid differentiation of monocytes into type I IFN-producing myeloid dendritic cells as an antiviral strategy against influenza virus infection. J Immunol. 2012;189(5):2257–65.

48. Hubo M, Trinschek B, Kryczanowsky F, Tuettenberg A, Steinbrink K, Jonuleit H. Costimulatory molecules on immunogenic versus tolerogenic human dendritic cells. Front Immunol. 2013;4:82.

49. Wijewardana V, Kristoff J, Xu C, Ma D, Haret-Richter G, Stock JL, et al. Kinetics of myeloid dendritic cell trafficking and activation: impact on progressive, nonprogressive and controlled SIV infections. PLoS Pathog. 2013;9(10):e1003600.

50. Sprenger HM, R.G.; Kaufmann, A.; Bussfeld, D.; Rischkowsky, E.; Gemsa, D. Selective induction of monocyte and not neutrophil-attracting chemokines after influenza A virus infection. J Exp Med. 1996;184:1191–6.

51. Gerlach RL, Camp JV, Chu YK, Jonsson CB. Early host responses of seasonal and pandemic influenza A viruses in primary well-differentiated human lung epithelial cells. PLoS One. 2013;8(11):e78912.

52. Dean RA, Cox JH, Bellac CL, Doucet A, Starr AE, Overall CM. Macrophage-specific metalloelastase (MMP-12) truncates and inactivates ELR+ CXC chemokines and generates CCL2, -7, -8, and -13 antagonists: potential role of the macrophage in terminating polymorphonuclear leukocyte influx. Blood. 2008;112(8):3455–64.

53. Tsou CL, Peters W, Si Y, Slaymaker S, Aslanian AM, Weisberg SP, et al. Critical roles for CCR2 and MCP-3 in monocyte mobilization from bone marrow and recruitment to inflammatory sites. J Clin Invest. 2007;117(4):902–9.

54. Molony RD, Nguyen JT, Kong Y, Montgomery RR, Shaw AC, Iwasaki A. Aging impairs both primary and secondary RIG-I signaling for interferon induction in human monocytes. Sci Signal. 2017;10.

55. Marr N, Wang TI, Kam SH, Hu YS, Sharma AA, Lam A, et al. Attenuation of respiratory syncytial virus-induced and RIG-I-dependent type I IFN responses in human neonates and very young children. J Immunol. 2014;192(3):948–57.

56. Strickland DH, Fear V, Shenton S, Wikstrom ME, Zosky G, Larcombe AN, et al. Persistent and compartmentalised disruption of dendritic cell subpopulations in the lung following influenza A virus infection. PLoS One. 2014;9(11):e111520.

57. Peruzzi B, Bencini S, Capone M, Mazzoni A, Maggi L, Salvati L, et al. Quantitative and qualitative alterations of circulating myeloid cells and plasmacytoid DC in SARS-CoV-2 infection. Immunology. 2020;161(4):345–53.

58. ACP. Normal Laboratory Values 2018. Available from: http://idgateway.wustl.edu/Normal%20lab%20values.pdf.

59. Charlson M, Pompei P, Ales K, MacKenzie C. A new method of classifying prognostic comorbidity in longitudinal studies- development and validation. J Chronic Dis. 1987;40(5):373–83.

60. Vincent JL, Moreno R, Takala J, Willatts S, De Mendonça A, Bruining H, et al. The SOFA (Sepsis-related Organ Failure Assessment) score to describe organ dysfunction:failure. On behalf of the Working Group on Sepsis-Related Problems of the European Society of Intensive Care Medicine. Intensive Care Med. 1996;22(7):770–10.

61. Grissom CK, Brown SM, Kuttler KG, Boltax JP, Jones J, Jephson AR, et al. A modified sequential organ failure assessment score for critical care triage. Disaster Med Public Health Prep. 2010;4(4):277–84.

62. Intensivvårdsregistret S. SIR:s riktlinje för registrering av SOFA. 2018 [Available from: https://www.icuregswe.org/globalassets/riktlinjer/sofa.pdf.

63. Svensson MJ, Lind I, Wirgart BZ, Ostlund MR, Albert J. Performance of the Simplexa Flu A/B & RSV Direct Kit on respiratory samples collected in saline solution. Scand J Infect Dis. 2014;46(12):825–31.

