## Supplementary table 1 for "Human influenza virus infection elicits distinct patterns of monocyte and dendritic cell mobilization in blood and the nasopharynx"

**Supplementary Table - Flow cytometry panels**

Panel 1: Phenotyping of MNPs from PBMCs

| Fluorochrome | Marker | Company | Clone |
|---|---|---|---|
| FITC | CD83 | Biolegend | HB15e |
| PerCp Cy5.5 | CD123 | BD | 7G3 |
| PE Cy7 | CD1c | Miltenyi | AD5-8E7 |
| PE Cy5 | CD11c | BD | B-Ly6 |
| PE TR | HLA-DR | Life Technologies | TU36 |
| PE | CD141 | Miltenyi | AD5-14H12 |
| APC-Cy7 | CD3 | BD | SK7 |
| | CD19 | BioLegend | HIB19 |
| | CD20 | BD | L27 |
| | CD56 | BD | HCD56 |
| | CD66abce | Miltenyi | TET2 |
| AF700 | CD16 | BioLegend | 3GE |
| APC | CD45 | BD | HI30 |
| BV786 | CD62L | BD | SK11 |
| BV650 | CD86 | BD | 2331 |
| BV605 | CCR2 | BioLegend | K036C2 |
| BV510 | CD14 | BD | M5E2 |
| V450 | CCR7 | BD | 150503 |
| DAPI | Live/Dead Blue | Thermo Fisher | Cat no: L34962 |

Panel 2: Phenotyping of PBMCs following TLR stimulation- TNF-release assay

| Fluorochrome | Marker | Company | Clone |
|---|---|---|---|
| FITC | CD83 | Biolegend | HB15e |
| PerCp Cy5.5 | CD123 | BD | 7G3 |
| PE Cy7 | CD1c | Miltenyi | AD5-8E7 |
| PE Cy5 | CD11c | BD | B-Ly6 |
| PE TR | HLA-DR | Life Technologies | TU36 |
| PE | **TNF-α Detection Antibody** | Miltenyi | (Reagent from 130-091-268) |
| APC-Cy7 | CD3 | BD | SK7 |
| | CD19 | BioLegend | HIB19 |
| | CD20 | BD | L27 |
| | CD56 | BD | HCD56 |
| | CD66abce | Miltenyi | TET2 |
| AF700 | CD16 | BioLegend | 3GE |
| APC | CD141 | Miltenyi | AD5-14H12 |
| BV786 | CD62L | BD | SK11 |
| BV650 | CD86 | BD | 2331 |
| BV605 | CCR2 | BioLegend | K036C2 |
| BV510 | CD14 | BD | M5E2 |
| V450 | CCR7 | BD | 150503 |
| DAPI | Live/Dead Blue | Thermo Fisher | Cat no: L34962 |
